# High-Dimensional Heterogeneity in Heat-Related Health Impacts Across German Districts

**DOI:** 10.64898/2026.09.23.26363662

**Authors:** Maria Gueltzow, Tarik Benmarhnia, Marcus Ebeling, Christopher Irrgang

## Abstract

**Background:** Multiple factors can drive heat susceptibility but are typically analyzed individually. This study investigates high-dimensional heterogeneity across multiple domains in the effect of hot days on emergency hospitalizations across German districts.

**Methods:** We obtained emergency hospitalization rates per 1,000 in 3-day windows for the summers of 2005-2023 from the German Diagnosis-Related Groups-Statistic, linked with spatio-temporal temperature information. A hot day was defined as at least one day above the district-level 98th percentile heat index. Multiple characteristics of the demographic, health/social, and environmental domains were selected as effect modifiers. We implemented a generalized random forest, where outcome and propensity scores were modeled with fixed effects estimators to control for seasonality and district-level confounding. We divided the conditional average treatment effects into quartiles to map their spatial distribution and identify key effect modifiers via the variable importance and observed indicator profiles across quartiles.

**Results:** While the average effect of heat on emergency hospitalizations was close to null (-0.3 per 1,000), effects in the lowest and highest effect quartiles ranged from -2.3 to +1.4 per 1,000. Demographic indicators explained the largest share of heterogeneity (34.5%), followed by environmental (14.53%) and health/-social factors (10.28%). The indicator profile for the most susceptible districts was characterized by lower air pollution levels, a larger percentage of vulnerable populations, and higher car density.

**Conclusion:** We highlight that heat susceptibility cannot be summarized by one number per region or be explained by a single effect modifier, which needs to be considered in heat-health warning systems and adaptation planning.

## Introduction

The increasing occurrence of hot days in a changing climate presents a major public health concern across the world [1]. This is exemplified by the reported impact of extreme heat on heat-related illness, mortality, and the worsening of chronic conditions [2, 3]. Besides the significant burden on population health, these potential health effects strain the health care system during periods of extreme heat through an increased number of unplanned and emergency care visits [4].

However, the impacts of heat are not distributed equally across communities. A variety of different factors were suggested in previous literature as potential drivers of this differential susceptibility to heat [5, 6]. Commonly, these include sociodemographic factors like age [4], race or ethnicity [7], or socioeconomic status [6]. Furthermore, health-related factors, such as pre-existing health conditions and medication usage [8], and factors related to the natural and built environment, such as air pollution levels [9] and the degree of urbanization [7], were proposed before. Most of the existing epidemiological literature typically focus on selected effect modifiers and analyze them separately [5, 6]. Yet, susceptibility to heat does not operate through isolated characteristics but through their intersection, as populations are simultaneously exposed to overlapping demographic, socioeconomic, and environmental disadvantages [10, 11].

Insights on which communities are most susceptible and identifying the potential drivers of heterogeneity are needed for health care planning and informing adaptation strategies in the context of climate change. While traditionally used methods such as stratified analyses or two-stage approaches with a meta-regression framework [5, 6] do not allow for the joint quantification of multiple effect modifiers, generalized random forests (grf) are well suited to this task. For example, Letellier et al. [12] recently used a grf to construct vulnerability profiles for adverse health outcomes following wildfire smoke exposure in California, jointly considering a large set of individual- and area-level modifiers. In this paper, we apply the grf framework in a spatio-temporal setting [13] to assess the effect of hot days on emergency hospitalizations across German districts, and aim to identify the most important regional drivers of effect heterogeneity. To the best of our knowledge, this is the first study to extend this high-dimensional, causal-forest–based framework to heat-related hospitalizations, accounting for the joint, intersectional structure of the candidate indicators.

## Methods

### Emergency Hospitalizations

We used the Diagnosis-Related Groups Statistic (DRG) to obtained information on all fully stationary hospitalizations in Germany from 2005-2023 that were accounted for by the DRG case rates [14]. Due to data protection reasons, we analyzed emergency hospitalizations aggregated within three day intervals and by administrative districts. We excluded cases from patients living abroad (0.6%), and with no information on age (*<* 0.01%) or sex/gender (*<* 0.01%). Additionally, 0.06% of all rows were removed because of observations below four cases. This resulted in 136,629,645 emergency hospitalizations, of which 34,376,508 occurred during summer (June to August). To account for changes in legislation in the definition of districts over the time span of our study, we harmonized the districts to year 2023 resulting in 400 districts.

We calculated the emergency hospitalization rate per 1,000 individuals by district and 3-day window based on the population size of each district, assuming that the population size is constant in a given year [15]. Population size was divided by 121 (the total number of 3-day categories in a year) to obtain the approximate person-years during a 3-day interval.

### Meteorological Data

We collected hourly information on mean temperature and the dew point on a 1 × 1 km grid from the HOSTRADA database, provided by the German Weather Service (DWD) [16]. This information was spatially matched with Census population data at the same resolution [17]. For each district, we computed hourly population-weighted averages of the weather variables across all grid cells within each district. The resulting hourly district-level information was subsequently aggregated to daily values.

We calculated a heat index with the mean daily temperature and dew point based on the Heat Index Equation of the National Weather Service [18]. In our main analysis, a heat event was defined as the heat index surpassing the 98th percentile of the summer daily heat index for at least one day within the 3-day window in a given district. The percentile cut-offs were calculated separately for each district based on the summer months during the reference period 1961-1990 (supplementary Figure S3).

### Measures of Heterogeneity

We screened previous literature to select relevant regional indicators for heat susceptibility. We identified 27 indicators, grouped into the demographic, health and social, and natural and built environment domains. We obtained information on these indicators from the INKAR database [19] which had sufficient information on 19 out of 27 indicators. If multiple variables from INKAR matched an identified heterogeneity indicator, we selected the ones that could cover different aspects of a domain (see Table S2). Missing information of indicators were filled in with the value of the previous or (if not available) following year. Information on monthly levels of air pollution (PM2.5) on a 9 × 9 km grid was obtained from the Atmospheric Composition Analysis Group [20]. To avoid multicollinearity issues in our analysis, we excluded strongly correlated features (see Figure S1). The final selection of indicators and their distribution is shown in Table 1. Additional information on the identification of indicators can be found in supplement section 6.

**Table 1:**
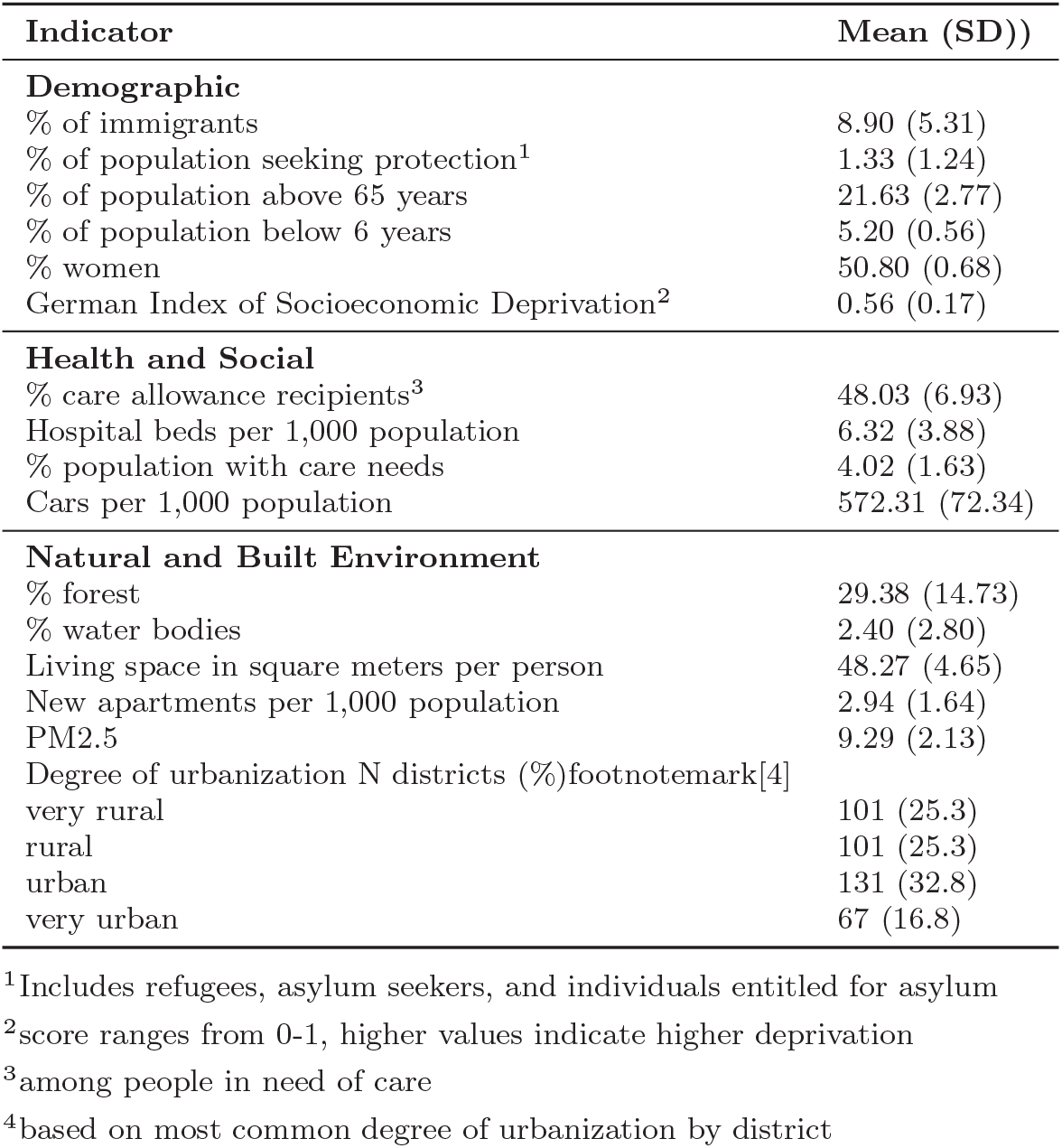
Selected Heterogeneity Indicators. Value ranges are indicated as mean values and standard deviations (SD).

| **Indicator** | **Mean (SD))** |
|---|---|
| **Demographic** |  |
| % of immigrants | 8.90 (5.31) |
| % of population seeking protection<sup>1</sup> | 1.33 (1.24) |
| % of population above 65 years | 21.63 (2.77) |
| % of population below 6 years | 5.20 (0.56) |
| % women | 50.80 (0.68) |
| German Index of Socioeconomic Deprivation<sup>2</sup> | 0.56 (0.17) |
| **Health and Social** |  |
| % care allowance recipients<sup>3</sup> | 48.03 (6.93) |
| Hospital beds per 1,000 population | 6.32 (3.88) |
| % population with care needs | 4.02 (1.63) |
| Cars per 1,000 population | 572.31 (72.34) |
| **Natural and Built Environment** |  |
| % forest | 29.38 (14.73) |
| % water bodies | 2.40 (2.80) |
| Living space in square meters per person | 48.27 (4.65) |
| New apartments per 1,000 population | 2.94 (1.64) |
| PM2.5 | 9.29 (2.13) |
| Degree of urbanization N districts (%)footnotemark[4] |  |
| very rural | 101 (25.3) |
| rural | 101 (25.3) |
| urban | 131 (32.8) |
| very urban | 67 (16.8) |

<sup>1</sup>Includes refugees, asylum seekers, and individuals entitled for asylum
<sup>2</sup>score ranges from 0-1, higher values indicate higher deprivation
<sup>3</sup>among people in need of care
<sup>4</sup>based on most common degree of urbanization by district

### Statistical Analysis

Our estimand of interest is the Conditional Average Treatment Effect (CATE) which is defined as

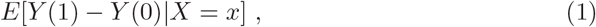

where *Y* (1) and *Y* (0) refer to the expected emergency hospitalization rate in the case of a heat event and no heat event, conditional on subgroup *X* = *x*.

We quantify the CATE and effect heterogeneity with a generalized random forest (grf) using the grf R package [21]. The methodological workflow can be found in Figure S2. As input, we include the emergency hospitalization rate (outcome), the heat event indicator (treatment), and the heterogeneity indicators. Second, we fit two nuisance models to predict the outcome and obtain the propensity score for a heat event to address confounding. Our analytical approach to address confounding can be seen as an extension of case-crossover designs in which temporal confounding is addressed using fixed effects models [22]. Specifically, we fit the outcome and propensity score model using fixed effects Poisson and logit estimators, with fixed effects for district, year and month to capture both long- and short-term seasonal trends. Standard errors were clustered at the district level. The model specifications can be found in the supplemental section 6.

In the third step, we included the predicted emergency hospitalization rate per 1,000 inhabitants and the propensity score as additional input alongside the observed data into a causal forest. The causal forest was clustered at the district-level, run with 10,000 trees, a minimum node size of 50 and alpha = 0.05. Other parameters were tuned using cross-validation to achieve the lowest predicted debiased error. We obtained the double-robust CATEs using augmented inverse probability weighting (AIPW) based on the outcome prediction and propensity score. To avoid extreme weights driven by the low prevalence of our exposure (13.1%), we used stabilized weights to construct the scores.

To gain deeper insights into the treatment effect heterogeneity within and across districts, we fit a second grf to rank each 3-day category into quartiles according to their estimated CATE prediction. We obtain the quartile CATE estimates by averaging the double-robust scores and assign each district its most commonly occurring CATE quartile. The percentage of days falling in a respective CATE quartile are shown in Figure S8. We furthermore obtain the leave-one-out treatment effect variable importance metric (TE-VIM) based on Hines et al. [23] by indicator and by domain. The scaled TE-VIM describes the proportion of the treatment effect heterogeneity explained by a respective indicator. To assess whether heterogeneity indicator profiles differ across CATE quartiles, we obtain the average values within each quartile for the 10 most important indicators. Since responses of the health care system to heat may not be immediate, we present the results for the heat event window (lag 0, days 0-2), short-term lag (lag 1, days 3-5), and extended lag (lag 2, days 6-8) to illustrate the delayed impact of heat of up to one week.

### Sensitivity Analysis

The definition of a heat day differs largely across the literature. We repeated our analysis with an absolute cut-off of 24°C average heat index for at least one day, and a heatwave definition of at least three days of a heat index above the 98th percentile in the current or previous 3-day category. To see whether our findings are affected by the Covid-19 pandemic, we exclude the years 2020-2023.

## Results

### Sample Characteristics

We observe 34,376,508 emergency hospitalizations in the summer months of 2005-2023. This translates to an average of 90.69 (SD: 26.29) emergency hospitalizations per 1,000 per district and 3-day window. The average local district-level heat index cut-off is 23.69 °C (SD:1.04), with a heat event prevalence of 13.1% during the summer months.

### Effect of a Heat Event on Emergency Hospitalizations across and within Districts

On average, a district experiencing at least one heat day within a 3-day window in the summer has a reduction of 0.3 (95%CI: -0.36;-0.24) per 1,000 (or 3 cases less per 10,000) in the emergency hospitalization rate. For the delayed effects, we find a decrease in the emergency hospitalization rate of 0.03 (95%CI: -0.05; -0.001) on days 3-5 and of 0.16 (95%CI: -0.18;-0.13) per 1,000 on days 6-8 since a heat event (Table S4).

Figure 1 shows the individual CATE predictions by district for the immediate effect (lag 0) of a heat event. For most districts, the mean and median of the individual CATE predictions are centered around zero or negative except for some districts in the South. However, we observe substantial variation across CATE predictions within a district, ranging between an increase and a decrease in the emergency hospitalization rate given a heat event, except for Eastern districts where CATE predictions were mostly negative. In contrast, the CATE predictions for the delayed response mostly show an increase in the emergency hospitalization rate on days 3-5 since a heat event for those Eastern districts Figure S5.

**Fig 1:**
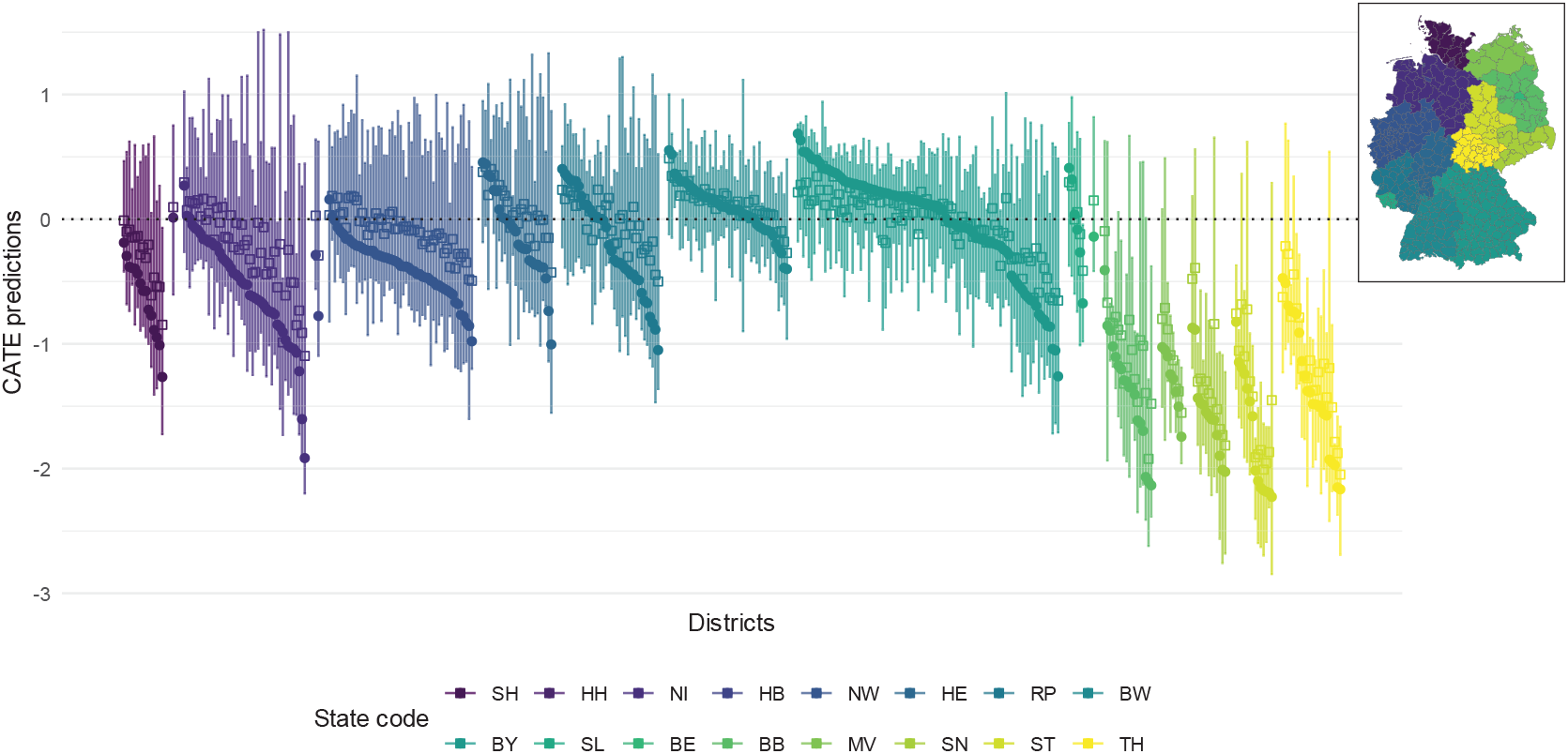
Individual CATE predictions for the immediate effect (days 0-2) of a heat event on the emergency hospitalization rate per 1,000, by district and grouped by state. Squares represent the mean, points the median. Bars represent the 25th to 75th percentile ranges. CATE predictions for the delayed effects (days 3-8) can be found in supplement Figure S5

To allow for more insights into the effect heterogeneity across districts, we divided the CATE estimates into quartiles. On average, we find a decrease in the emergency hospitalization rate by -2.3 (95%CI: -2.4;-2.2) and -0.66 (95%CI: -0.7;-0.62) per 1,000 for 3-day categories falling into Q1 (moderate decrease) and Q2 (minor decrease), and increases by 0.27 (95%CI: 0.24; 0.31) and 1.41 (95%CI: 1.36; 1.46) per 1,000 for 3-day categories falling into Q3 (minor increase) and Q4 (moderate increase). The quartile CATE estimates for the delayed effects are comparable to the immediate response (days 0-2), with a larger CATE for the moderate decrease quartile on days 3-5 since a heat event(Figure S7).

Figure 2 shows the most common CATE quartile per district. The North Eastern districts most commonly experience an immediate moderate decrease, and the Southern districts most commonly experience a minor to moderate increase in the emergency hospitalization rate within a heat event window. The Western districts show more heterogeneity ranging from a minor decrease to a moderate increase. The delayed response follows a different pattern, with an increase in the emergency hospitalization rate days 3-5 since a heat event mainly in the East and North West of Germany, and a moderate increase in the emergency hospitalization rate days 6-8 since a heat event mainly in the Central Northern regions.

**Fig 2:**
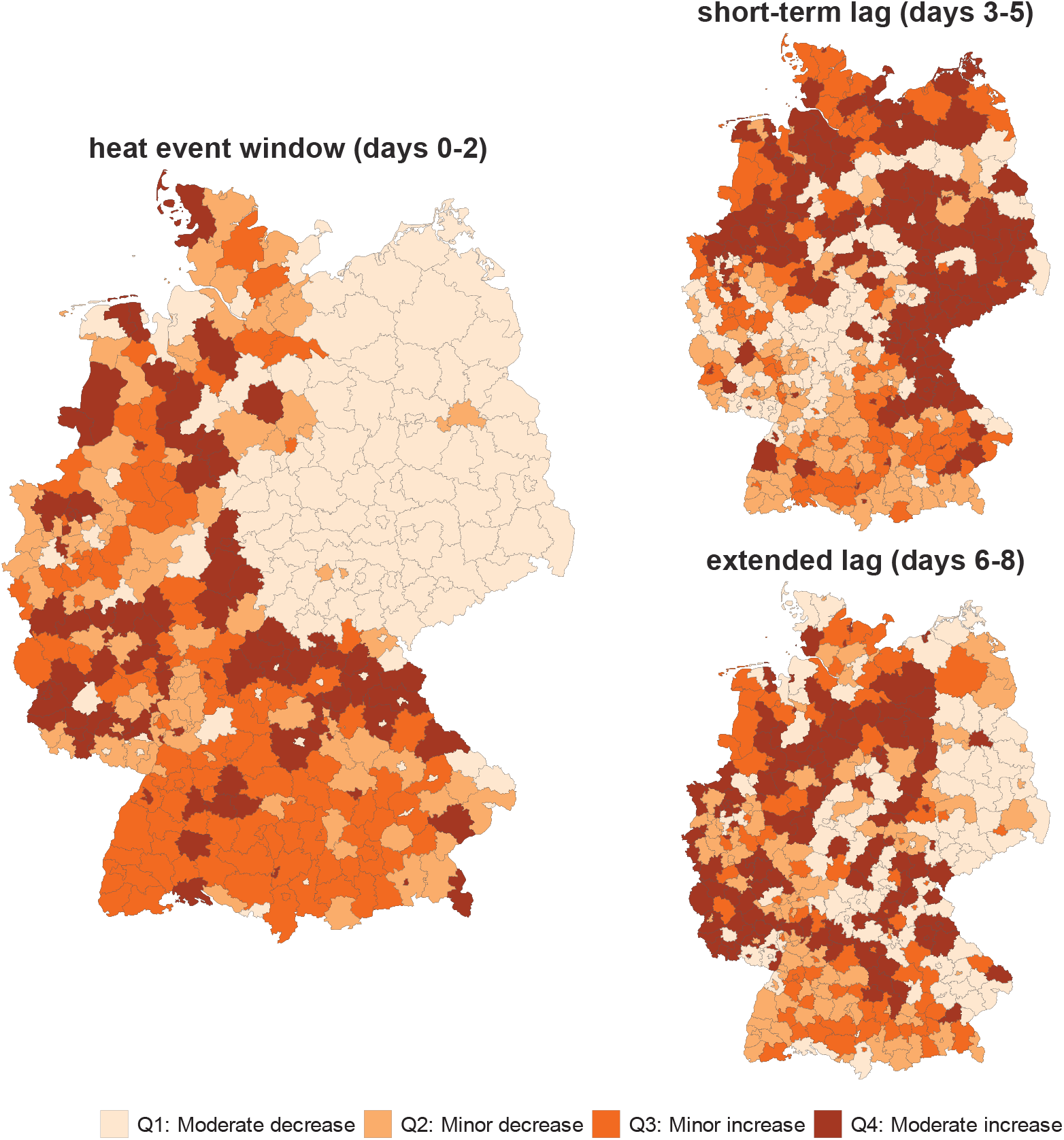
Most common quartile of the effect of a heat event on emergency hospitalizations by district across lags.

### Drivers of Heat Susceptibility

The calibration output indicates that the causal forest successfully detected effect heterogeneity (Table S3. Overall, the demographic domain explains 34.5% (95% CI: 32.37; 36.73) of the effect heterogeneity in the heat-emergency hospitalization relationship, followed by 14.53% (95% CI: 13.03; 16.02) for the environmental domain, and 10.28% (95% CI: 8.97; 11.59) for the health & social domain. Figure 3 shows that for the demographic indicators, the proportion of people seeking protection was the most important variable for predicting heterogeneity in the effect of heat on the emergency hospitalization rate. This was followed by the percentage of people above 65 years, the percentage of migrants, percentage of people under six years, and percent-age of women. In the health and social domain, car density per 1,000, percentage of population with care needs and the population receiving care allowance were the most important for predicting treatment effect heterogeneity. For the environmental indicators, the particular matter (PM2.5) and living space in square meters per person were the most important.

**Fig 3:**
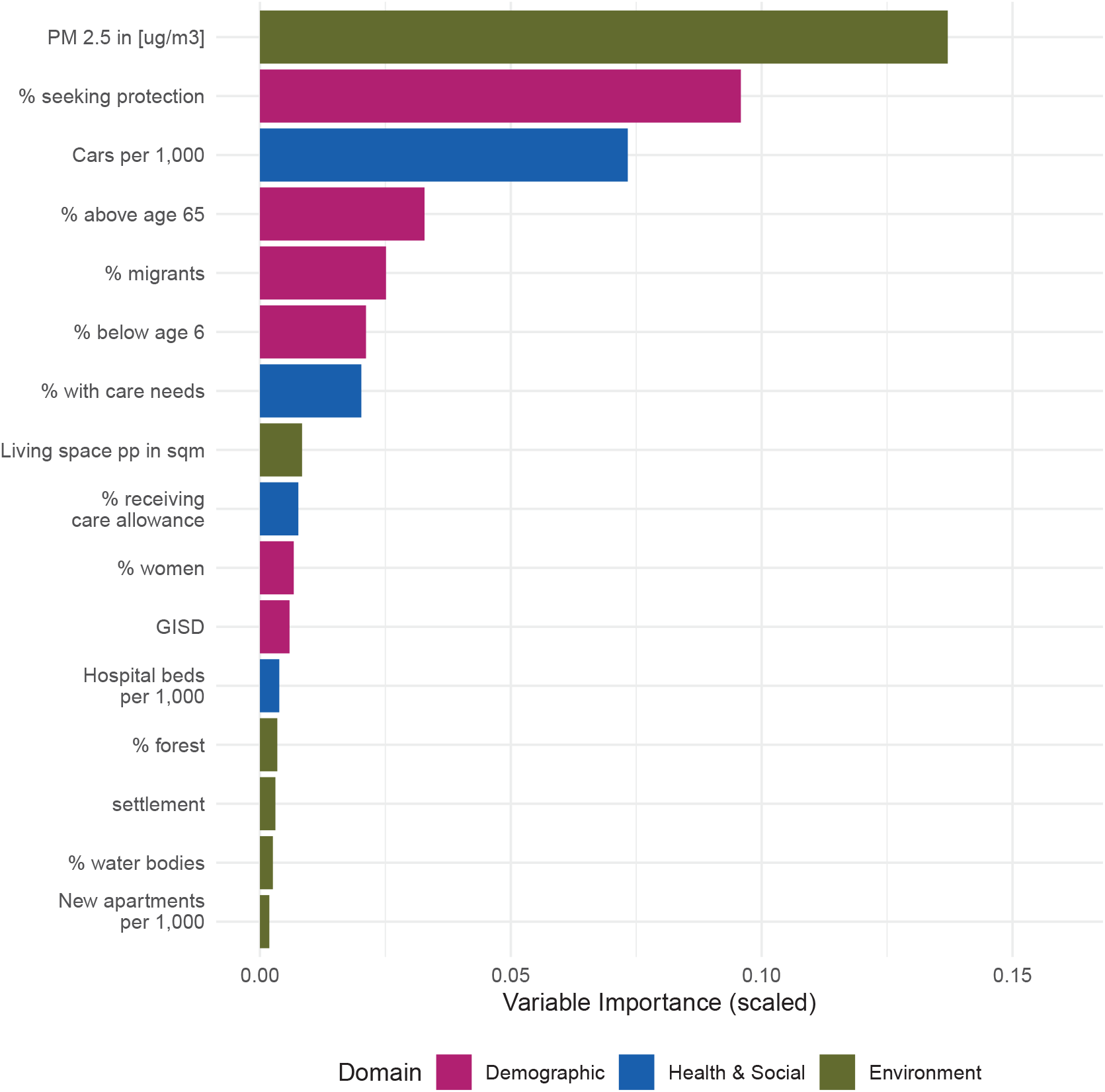
Variable importance (scaled) for predicting effect heterogeneity in the immediate effect (day 0-2) of a heat event on the emergency hospitalization rate by domain.

A larger percentage of the effect heterogeneity can be explained by the selected indicators for the delayed response with more than 50% of effect heterogeneity explained by air pollution on days 3-5 and 6-8 since a heat event. Otherwise similar indicators were deemed important (Figure S6).

The average indicator values within each heat effect quartile Figure 4 reveal a gradient across quartile groups with a larger average percentage of people seeking protection (Q4: 2.2%; Q1: 0.8%), percentage of migrants (Q4: 11%; Q1: 5.3%), percentage receiving care allowance (Q4: 50%; Q1: 46%), car density (Q4: 600 per 1,000; Q1: 550 per 1,000), and living space (Q4: 50 sqm; Q1: 47 sqm), but lower air pollution levels (Q4: 8.3 ug/m3; Q1: 10 ug/m3) among the moderate increase group (Q4). The percentage of older adults (Q1: 23%; Q4: 22%) and the percentage with care needs (Q1: 4.7%; Q4: 5.5%) is largest in both the moderate decrease and moderate increase heat effect quartiles, while the percentage of children is larger in the minor (5.4%) and moderate increase quartile (5.5%).

**Fig 4:**
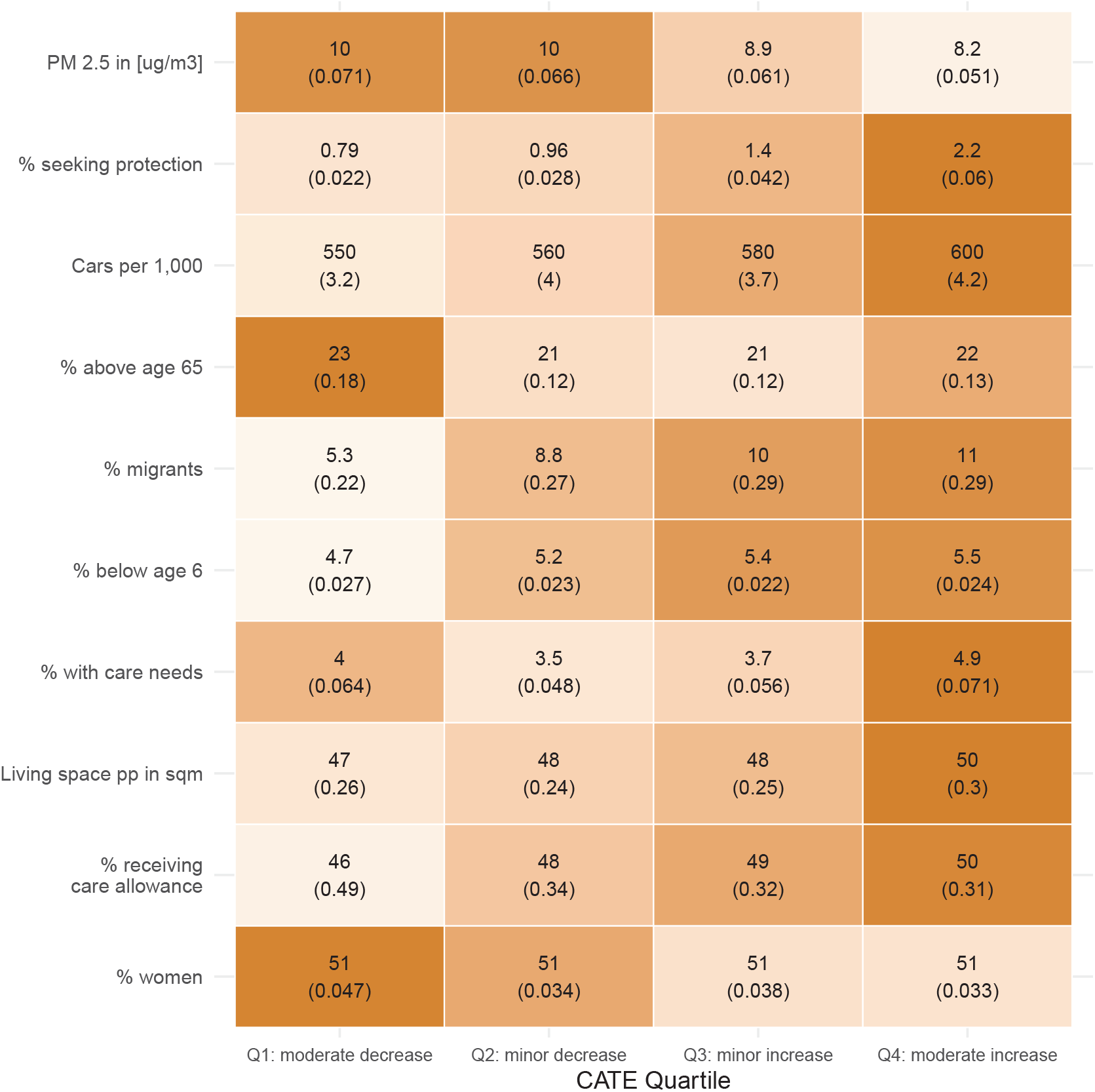
Average indicator values within heat effect quartiles. Results are shown for the top 10 most important variables according to variable importance metric. The heat effect describes the immediate effect (days 0-2) of a heat event on the emergency hospitalization rate. Number in brackets shows the standard error. Darker colors indicate a larger average value of an indicator in a given quartile, in comparison to other quartiles.

Compared to the immediate response (days 0-2), we find a reversed trend in average indicator values across heat effect quartiles on days 3-5 since a heat event for most indicators, with the larger air pollution levels among the moderate increase quartile (Q4: 9.8%; Q1: 8.9%) but a lower percentage of people with care needs (Q4: 3.8%; Q1: 5.1%), migrants (Q4: 6.7%; Q1: 9.6%), population seeking protection (Q4: 1%; Q1: 1.9%), car density (Q4: 560; Q1: 590 per 1,000) and population receiving care allowance (Q4: 46%; Q1: 50%) compared to the immediate response. Trends for days 6-8 since a heat event are less clear (Figure S9, Figure S10).

### Sensitivity Analyses

We performed robustness checks for alternative heat definitions and excluding the Covid-19 years. While results for the absolute heat cut-off were comparable to the main analysis, excluding years 2020-2023 and using the heatwave definition resulted in the percentage of people with care needs being identified as one of the most important predictors. The heatwave cut-off also showed a higher CATE and a larger percentage in treatment effect heterogeneity explained by the selected indicators. Otherwise results were comparable. A more detailed description of the findings can be found in the supplement section 6.

## Discussion

We investigated how the effect of hot days on emergency hospitalizations varies across and within German districts and how regional indicators, spanning the demographic, health and social, and environmental domains, jointly shape the heterogeneity in this effect. We found that on average across all districts, a heat event resulted in a modest reduction of 3 emergency hospitalizations per 10,000 persons (95%CI: -3.6; -2.4). However, the effect showed a considerable regional variability with districts located in the South and West mostly experiencing a minor to moderate increase in the emergency hospitalization rate immediately following heat (days 0-2), whereas the East and North West of Germany experienced increases on days 3-5 since a heat event. The effect heterogeneity was mostly explained by demographic indicators, followed by indicators of the environmental and health & social domain, with air pollution levels, the percentages of the population seeking protection, and car density as the most important individual indicators.

We found substantial variation in the estimated effect of heat within districts, not only across them. For most districts the individual heat effect predictions spanned both increases and decreases in the emergency hospitalization rate. Most previous heat–health research characterized spatial variability in a largely static manner by estimating one summary risk per area. A growing body of studies advanced this approach by mapping temperature-related health risks at a fine spatial resolution [3, 24–27] and demonstrated that heat-related health risks are not only socially patterned but also spatially uneven. This study complements this literature by making the within-area variability the study objective. Our findings indicate that treating the heat–health relationship within a given geographical area as static across time does not capture how emergency hospitalizations actually respond to heat over the study period. Therefore, approaches that assign each region a single risk estimate cannot, by construction, represent its internal variation. Our approach provides this more granular information and situates it within a coherent estimand.

Apart from the heterogeneity within districts, we also found a clear spatial gradient across districts which differs across time. Districts in the West most often showed substantial variation with moderate decreases to increases in the emergency hospitalization rate immediately following heat (days 0-2), while districts in the South mostly experienced an increase. However, in the East, the immediate effect of heat mainly led to a moderate decrease in the emergency hospitalization rate but a moderate increase on days 3-5 since the heat event. This gradient persists in simple Poisson models (S20) and after excluding indicators with a pronounced East-West gradient (S21) though the pattern showed a broader regional gradient rather than the former inner-German border. Frasch et al.[28] and Kriit et al. [29] also found a negative association between extreme heat and emergency hospitalizations for some Eastern regions in Germany. Districts in the East of Germany tend have larger distances to the next hospital [30] which might lead to hesitancy to seek emergency medical services in situations that do not present clear-cut emergencies. The observed trend therefore might represent different healthcare system structures, delayed response times or different care seeking behavior in the East compared to other regions. Future research is needed to further investigate this finding.

We find that heat susceptibility is shaped jointly by indicators from distinct domains rather than by a single indicator alone. Overall, effect heterogeneity in the heat-hospitalization relationship was mainly driven by demographic and environmental factors, whereas the health and social domain was important mainly for predicting effect heterogeneity in delayed effects. The distribution in demographic indicators across heat effect quartiles showed that a moderate increase in the emergency hospitalization rate following heat occurs in districts with a larger percentage of people seeking protection, migrants, population with care needs, children and older adults. This indicates that districts with the most vulnerable populations show higher heat-related emergency hospitalizations which aligns with previous findings (e.g., [4, 6, 7]).

Additionally, our results regarding indicators of the environmental, and health and social domain point to districts with higher car density, larger average living space, and lower air pollution to be particularly susceptible. While air pollution was so far described as increasing the detrimental effect of heat on health overall [31], we find this to be the case only for the delayed effects of heat. Furthermore, higher car density and higher living space are negatively correlated with population size (supplementary Figure S1), pointing to more sparsely populated districts with lower access to hospital care being susceptible to heat.

Several limitations should be considered when interpreting our findings. First, for data-protection reasons the hospitalization data were available only in three-day windows. This temporal resolution might miss day-to-day fluctuations in emergency hospitalizations due to heat; nonetheless, the three-day window still captures the main acute effects of heat on emergency care, which are known to occur over a short lag. Second, our main exposure was defined using a heat index that combines temperature and dew point, and we did not examine the full range of alternative heat metrics, including other humidity-based measures. We addressed this in part through sensitivity analyses using an absolute cut-off and a heatwave definition, which produced broadly consistent results. A more comprehensive comparison of heat metrics remains a natural extension for future work. Third, although we screened the literature systematically for relevant indicators spanning three domains, future studies could expand the heterogeneity indicator set. This includes structural indicators of the health care system, such as distance to nearest doctor or hospital bed occupancy, as well as other climatic indicators, like solar radiation or the time since last heat event. Finally, our approach clusters at the district level but does not explicitly capitalize on the spatial structure of the data, and residual spatial autocorrelation may remain. Future extensions could embed the framework within a Bayesian hierarchical spatial model to borrow strength across neighboring districts and improve statistical precision, as was done in related small-area and multi-scale heat–health analyses [26, 27].

## Conclusion

There is substantial variation in heat susceptibility across and within districts in Germany, and this variation is driven by the interplay of indicators from the demographic, health and social, and natural and built environment domains. Summarizing heat impacts through a single average, or through effect modifiers examined one at a time, does not capture this complexity. The grf quantifies the heterogeneity in the effect of heat on emergency hospitalizations and identifies its most important regional drivers while accounting for their joint structure. Understanding heat susceptibility as the joint, intersectional product of multiple district characteristics has direct relevance for heat action plans and heat–health warning systems. Warning systems and adaptation measures are frequently designed around generic categories of vulnerable groups or around a single dominant risk factor, which risks overlooking places where several moderate disadvantages combine to produce high susceptibility. Applied here to Germany, our approach offers a transferable way of documenting local, high-dimensional evidence on heat susceptibility that is needed to inform adaptation strategies and heat–health warning systems in a changing climate.

## Data Availability

This study uses restricted data from the G-DRG Statistic
(https://doi.org/10.21242/23141.2005.00.00.1.1.0 to https://doi.org/10.21242/23141.2023.00.00.1.1.0) through remote execution. Remote execution restricts users to submitting code for execution on the RDC servers and only aggregated results, subject to statistical disclosure control, are returned to the researcher. Data access can
be applied for through the Research Data Center (RDC) of the Federal Statistical Office and Offices (see https://www.forschungsdatenzentrum.de/en/request). All other data sources used in this study are publicly available. Population counts can
be obtained from https://www.destatis.de/EN, temperature information is available at https://www.dwd.de, the German Index of Socioeconomic Deprivation can be found on https://robert-koch-institut.github.io/German_Index_of_Socioeconomic_Deprivation_GISD/ and regional characteristics were obtained from https://www.inkar.de/.

## Acknowledgements

The authors would like to express their gratitude to Junyu Wang for the pre-processing of parts of the environmental data used in this study.

## Author contributions

MG: Conceptualization, Formal analysis, Investigation, Methodology, Project administration, Validation, Visualization, Writing – original draft, Writing – review & editing; TB: Conceptualization, Investigation, Methodology, Validation, Writing – original draft, Writing – review & editing; ME: Conceptualization, Methodology, Validation, Visualization, Writing – review & editing; CI: Conceptualization, Methodology, Validation, Visualization, Writing – review & editing

## Data Availability

This study uses restricted data from the G-DRG Statistic (https://doi.org/10.21242/23141.2005.00.00.1.1.0 to https://doi.org/10.21242/23141. 2023.00.00.1.1.0) through remote execution. Remote execution restricts users to submitting code for execution on the RDC’s servers and only aggregated results, subject to statistical disclosure control, are returned to the researcher. Data access can be applied for through the Research Data Center (RDC) of the Federal Statistical Office and Offices (see https://www.forschungsdatenzentrum.de/en/request). All other data sources used in this study are publicly available. Population counts can be obtained from https://www.destatis.de/EN, temperature information is available at https://www.dwd.de, the German Index of Socioeconomic Deprivation can be found on https://robert-koch-institut.github.io/German Index of Socioeconomic Deprivation GISD/ and regional characteristics were obtained from https://www.inkar.de/.

## Code Availability

All analyses were conducted using R (version 4.4.2). All code used to perform the analyses in this study is available at https://github.com/ClimSocAna/GRF-heat-hosp/.

## Declaration of generative AI use

During the preparation of this work, the authors used Claude Sonnet 4 to 5 for assistance with R code and for language editing of a limited number of sentences. No text, figures, or analytical output were generated by the tool and incorporated into the manuscript verbatim. All AI-assisted code and edits were reviewed and verified by the authors, who take full responsibility for the content of the publication.

## References

[1] Romanello M, Walawender M, Hsu SC, Moskeland A, Palmeiro-Silva Y, Scamman D, et al. The 2025 report of the Lancet Countdown on health and climate change: climate change action offers a lifeline. The Lancet. 2025 12;406(10521):2804–2857. 10.1016/S0140-6736(25)01919-1.

[2] Cissé G, McLeman R, Adams H, Aldunce P, Bowen K, Campbell-Lendrum D, et al. Health, Wellbeing, and the Changing Structure of Communities. In: Pörtner HO, Roberts DC, Tignor M, Poloczanska ES, Mintenbeck K, Alegría A, et al., editors. Climate Change 2022: Impacts, Adaptation and Vulnerability. Contribution of Working Group II to the Sixth Assessment Report of the Intergovernmental Panel on Climate Change. Cambridge, UK and New York, NY, USA: Cambridge University Press; 2022. p. 1041–1170.

[3] Wang J, Nikolaou N, ander Heiden M, Irrgang C. High-resolution modeling and projection of heat-related mortality in Germany under climate change. Communications Medicine. 2024 oct;4(1):1–8. 10.1038/s43856-024-00643-3.

[4] Li M, Gu S, Bi P, Yang J, Liu Q. Heat Waves and Morbidity: Current Knowledge and Further Direction-A Comprehensive Literature Review. International Journal of Environmental Research and Public Health. 2015 may;12(5):5256–5283. 10.3390/IJERPH120505256.

[5] Benmarhnia T, Deguen S, Kaufman JS, Smargiassi A. Vulnerability to heat-related mortality: A systematic review, meta-analysis, and meta-regression analysis. Epidemiology. 2015 oct;26(6):781–793. 10.1097/EDE.0000000000000375.

[6] Arsad FS, Hod R, Ahmad N, Ismail R, Mohamed N, Baharom M, et al. The Impact of Heatwaves on Mortality and Morbidity and the Associated Vulnerability Factors: A Systematic Review. International journal of environmental research and public health. 2022 12;19(23). 10.3390/IJERPH192316356.

[7] Gronlund CJ. Racial and socioeconomic disparities in heat-related health effects and their mechanisms: a review. Current epidemiology reports. 2014 9;1(3):165– 173. 10.1007/S40471-014-0014-4.

[8] Wu WJ, Hutton J, Zordan R, Ranse J, Crilly J, Tutticci N, et al. Review article: Scoping review of the characteristics and outcomes of adults presenting to the emergency department during heatwaves. Emergency medicine Australasia : EMA. 2023 12;35(6):903–920. 10.1111/1742-6723.14317.

[9] Ma Y, Chen C, Aguilera R, Gershunov A, Jerrett M, Connolly R, et al. Spatial heterogeneity in synergistic effects of extreme heat and NO2 exposures on cardiorespiratory hospitalizations in California. Epidemiology (Cambridge, Mass). 2026 3;10.1097/EDE.0000000000001970.

[10] Tschakert P, Ogra A, Sharma U, Karthikeyan K, Singh A, Bhowmik A. Intersecting inequalities and urban heat adaptation. Global Environmental Change. 2025 jul;92:103003. 10.1016/J.GLOENVCHA.2025.103003.

[11] Kc A, Kreyenbaum L. Global call to understand intersectionality between heat exposure and perinatal mental health. Maternal Health, Neonatology and Perinatology. 2025 mar;11(1):6. 10.1186/S40748-025-00206-X.

[12] Letellier N, Jones-Ngo CG, Cheung MW, Aguilera R, Casey JA, Zakaras JM, et al. Using generalized random forests to characterize vulnerability to adverse health outcomes following wildfire smoke exposure in California. Environment International. 2025 ec;206:109955. 10.1016/J.ENVINT.2025.109955.

[13] Athey S, Wager S. Estimating Treatment Effects with Causal Forests: An Application. Observational Studies. 2019;5(2):37–51. 10.1353/obs.2019.0001.

[14] RDC of the Federal Statistical Office and Offices. Diagnosis-Related Groups Statistic. RDC of the Federal Statistical Office and Offices; 2023. Available from: https://www.forschungsdatenzentrum.de/en/health/drg.

[15] Statistisches Bundesamt (Destatis). Population: Administrative Districts, Reference Date. Period: 31.12.2004 -31.12.2023. Statistisches Bundesamt (Destatis); 2025.

[16] German Weather Service (DWD).: HOSTRADA -Hochaufgeloester Stuendlicher Rasterdatensatz fuer Deutschland, Version 1.0. German Weather Service (DWD). Available from: https://www.dwd.de/DE/leistungen/uhi/infomethodic/01hostrada.html.

[17] Statistische Ä mter des Bundes und der Länder.: Ergebnisse des Zensus 2022: Bevölkerungszahlen in Gitterzellen. Statistisches Bundesamt.

[18] Rothfusz LP.: The Heat Index “Equation” (or, More Than You Ever Wanted to Know About Heat Index). Fort Worth, TX, US.

[19] Federal Institute for Building, Urban Affairs and Spatial Development (BBSR). Ongoing spatial observation by the BBSR – INKAR, issue 07/2025. Bonn: Federal Institute for Building, Urban Affairs and Spatial Development (BBSR); 2025. Available from: https://www.inkar.de/.

[20] Atmospheric Composition Analysis Group (ACAG).: Satellite-derived PM2.5 (V6.GL.02.04). Atmospheric Composition Analysis Group (ACAG). Available from: https://sites.wustl.edu/acag/surface-pm2-5/#V6.GL.02.04.

[21] Athey S, Tibshirani J, Wager S. Generalized random forests. Ann Statist. 2019;47(2):1148–1178. 10.1214/18-AOS1709.1610.01271.

[22] Tobias A, Kim Y, Madaniyazi L. Time-stratified case-crossover studies for aggregated data in environmental epidemiology: a tutorial. International Journal of Epidemiology. 2024 2;53(2). 10.1093/IJE/DYAE020.

[23] Hines OJ, Diaz-Ordaz K, Vansteelandt S. Variable importance measures for heterogeneous treatment effects. Biometrics. 2025 ec;81(4). 10.1093/biomtc/ujaf140.

[24] Gasparrini A, Masselot P, Scortichini M, Schneider R, Mistry MN, Sera F, et al. Small-area assessment of temperature-related mortality risks in England and Wales: a case time series analysis. The Lancet Planetary Health. 2022 jul;6(7):e557–e564. 10.1016/S2542-5196(22)00138-3.

[25] Masselot P, Mistry M, Vanoli J, Schneider R, Iungman T, Garcia-Leon D, et al. Excess mortality attributed to heat and cold: a health impact assessment study in 854 cities in Europe. The Lancet Planetary health. 2023 apr;7(4):e271–e281. 10.1016/S2542-5196(23)00023-2.

[26] Hansen K, Schwartzman A, Schwarz L, Teyton A, Basu R, Benmarhnia T. The spatial distribution of heat related hospitalizations and classification of the most dangerous heat events in California at a small-scale level. Environmental Research. 2024 nov;261:119667. 10.1016/J.ENVRES.2024.119667.

[27] Schwarz L, Chen C, Castillo Quiñones JE, Aguilar-Dodier LC, Hansen K, Sanchez JR, et al. Heat-related mortality in Mexico: A multi-scale spatial analysis of extreme heat effects and municipality-level vulnerability. Environment International. 2025 jan;195:109231. 10.1016/J.ENVINT.2024.109231.

[28] Frasch JJ, König HH, Konnopka C. Effects of extreme temperature on morbidity, mortality, and case severity in German emergency care. Environmental Research. 2025 apr;270:121021. 10.1016/J.ENVRES.2025.121021.

[29] Kriit HK, Herrmann A, Norris R, Rocklöv J, Allegri MD. Heat exposure and the risk of emergency hospitalization in Germany: Stratified analyses by age, sex, and diagnostic group. Deutsches Ä rzteblatt international. 2025 ec;10.3238/arztebl.m2025.0186.

[30] Statistische Ä mter des Bundes und der Länder. Krankenhausatlas. Statistische Ä mter des Bundes und der Länder; 2023. Available from: https://krankenhausatlas.statistikportal.de/.

[31] Patel D, Jian L, Xiao J, Jansz J, Yun G, Robertson A. Joint effect of heatwaves and air quality on emergency department attendances for vulnerable population in Perth, Western Australia, 2006 to 2015. Environmental research. 2019 7;174:80–87. 10.1016/J.ENVRES.2019.04.013.

