## Supplemental Material for "High-Dimensional Heterogeneity in Heat-Related Health Impacts Across German Districts"

### Supplementary Material

#### Additional Information on Indicator Selection

To identify relevant regional indicators for sensitivity to heat, we screened existing research on this topic published in PubMed and Web of Science from 2000 to March 2026. The search strategy can be found in supplement Table S1. We identified 196 papers from PubMed substituted with additional publications identified through Web of Science. We identified 41 relevant publications which reported on 27 unique indicators. Eight of these were categorized as indicators of the demographic domain, eight as indicators of the health and social domain, and 11 as indicators of the natural and built environment domain. Out of the identified indicators, five of them had no sufficient information available at the district level (homelessness, social cohesion/connectedness, medication use, soil sealing, practitioner density). The INKAR database did not provide information for homelessness, social cohesion/connectedness, medication use, soil sealing at the district-level. For practitioner density, information from the INKAR database contained implausible values and was therefore excluded from the list of selected indicators. Air Conditioning and health insurance coverage were excluded because they were not relevant in the German context as built-in air conditioning is not common in Germany, and health insurance coverage is mandatory. One identified indicators was humidity which is included in the heat index and was therefore excluded. This resulted in 19 indicators selected as potential drivers of effect heterogeneity (supplement Table S2).

**Table S1**: Literature search strategy to identify heterogeneity indicators.

| **Database** | **Search String** | **Additional information** | Date | Results |
|---|---|---|---|---|
| Pubmed | (”Hot Temperature”[MeSH] OR “heat”[tiab] OR ”high temperature”[tiab] OR ”heatwave*”[tiab] OR ”heat wave*”[tiab] OR ”extreme heat”[tiab] ) AND (”Hospitalization”[MeSH] OR ”Emergency Medical Services”[MeSH] OR “hospitalization”[tiab] OR ”hospital admission*”[tiab] OR ”emergency visit*”[tiab] OR ”healthcare utilization”[tiab]) AND (”Vulnerable Populations”[MeSH] OR ”Health Status Disparities”[MeSH] OR “vulnerability”[tiab] OR “susceptibility”[tiab] OR ”susceptible population*”[tiab] OR “heterogeneit*”[tiab] OR “inequalit*”[tiab] OR “inequit*”[tiab] OR “disparit*”[tiab] OR ”social gradient”[tiab] OR ”at-risk population*”[tiab] ) | Full screen of results | 09.03.2026 | 196 |
| Web of Science | TS=( heat OR ”high temperature” OR heatwave* OR ”heat wave*” OR ”extreme heat”) AND TS=(hospitalization OR ”hospital admission*” OR ”emergency visit*” OR ”healthcare utilization”) AND TS=(vulnerability OR susceptibility OR ”susceptible population*” OR heterogeneit* OR inequalit* OR inequit* OR disparit* OR ”social gradient” OR ”at-risk population*”) | Supplementary screen of additional publications not found in PubMed; Article or Review Article, English | 09.03.2026 | 360 |

**Table S2**: Identified heterogeneity indicators and availability for Germany based on INKAR database.

| Group | Indicator based on literature | Set of indicators available for Germany | Years available |
|---|---|---|---|
| Demographics | racial or ethnic minorities [1–10] | % migrants | 2005-2023 |
| | | % of people seeking protection[1] | 2007-2023 |
| | population density [2, 11, 12] | population per sq km | 2008-2023 |
| | old age [3, 5, 6, 10, 11, 13–22] | % of population above 65 years | 2005-2023 |
| | young age [3, 14, 15, 18, 23] | % population below 6 yeas | 2005-2023 |
| | gender [3, 4, 7, 15, 16, 18, 24] | % of women | 2005-2023 |
| | pregnancy [25, 26] | births per 1,000 population | 2005-2023 |
| | homeless [27] | *no information available* | - |
| | Socioeconomic deprivation [2, 3, 5–7, 9, 11–14, 16, 18, 24, 28–32] | German Socioeconomic Deprivation Index (GISD) | 2005-2023 |
| Health & Social | pre-existing health conditions [12, 15, 16, 18, 24, 28, 32–35] | life expectancy | 2005-2022 |
| | disability status [20, 36] | % of care allowance recipients | 2005-2022 |
| | health insurance coverage [2] | *not relevant in German context* | - |
| | automobile ownership [2] | cars per 1,000 population | 2005-2012, 2014-2023 |
| | social isolation/cohesion [30, 34] | *no information on aggregate level* | - |
| | medications [9, 16] | *no information on aggregate level* | - |
| | practitioner density [37] | *no sufficient information available* | - |
| | Number of hospitals [38] | Number of hospital beds per 1,000 | 2005-2022 |
| Natural & Built Environment | AC | *not relevant in German context* | - |
| | air pollution (PM2.5,SO2, NO2, O3) [3, 28, 39–41] | monthly levels of PM2.5[2] | 2005-2023 |
| | soil sealing [41] | *no information available* | - |
| | green space [38, 41] | % forest | 2016-2023 |
| | | % water bodies | 2016-2023 |
| | | % open space | 2016-2023 |
| | road and traffic density [41] | cars per 1,000 population | 2005-2012, 2014-2023 |
| | tree coverage [2, 28] | % forest | 2016-2023 |
| | small dwelling [29] | Average sqm of living space pp | 2011-2023 |
| | quality of homes [24, 42] | Number of new apartments per 1,000 population | 2006-2009, 2011, 2016-2023 |
| | urbanicity [9, 15, 24, 43] | Degree of urbanization (very rural, rural, urban, very urban) | 2017-2023 |
| | retirement homes [24] | % people with care needs | 2005-2022 |
| | humidity [10, 22] | daily dew point temperature[3] | 2005-2023 |

[1]includes refugees, asylum seekers, individuals entitled to asylum

[2]available from ACAG [44]

[3]available from HOSTRADA [45]

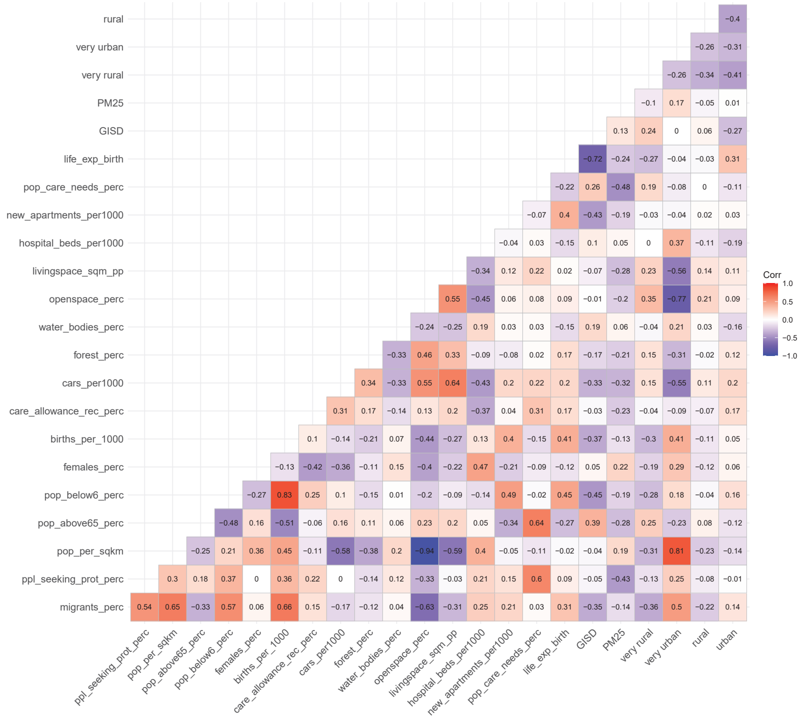

**Fig. S1**: Correlation matrix for heterogeneity indicators. Indicators with a correlation of above 0.7 were excluded to avoid issues with multicollinearity.

#### Methodological workflow and model diagnostics.

##### Fixed effects model specifications.

We determined the appropriate fixed effects structure based on the lowest BIC (Bayesian Information Criterion). The outcome model was defined as

$$\log(Y_{r,t}) = \beta_1 W_{r,t} + \alpha_{r,y} + \gamma_t + \log(\text{population}_{r,t}) \ , \tag{2}$$

where $Y_{r,t}$ are emergency hospitalizations per district and 3-day window, $W_{r,t}$ is a binary indicator of heat event per district and 3-day window, $\alpha_{r,y}$ defines district-by-year fixed effects, $\gamma_m$ fixed effects by month, and $\log(\text{population}_{r,t})$ defines a population offset.

The propensity score model was defined as

$$\log(W_{r,t}) = 1 + \alpha_r + \delta_y + \gamma_t \ , \tag{3}$$

where $\alpha_r$, $\delta_y$, and $\gamma_t$ define fixed effects for district, year, and month separately. The propensity score resulted in sufficient overlap between the exposed and unexposed groups (see Figure S4).

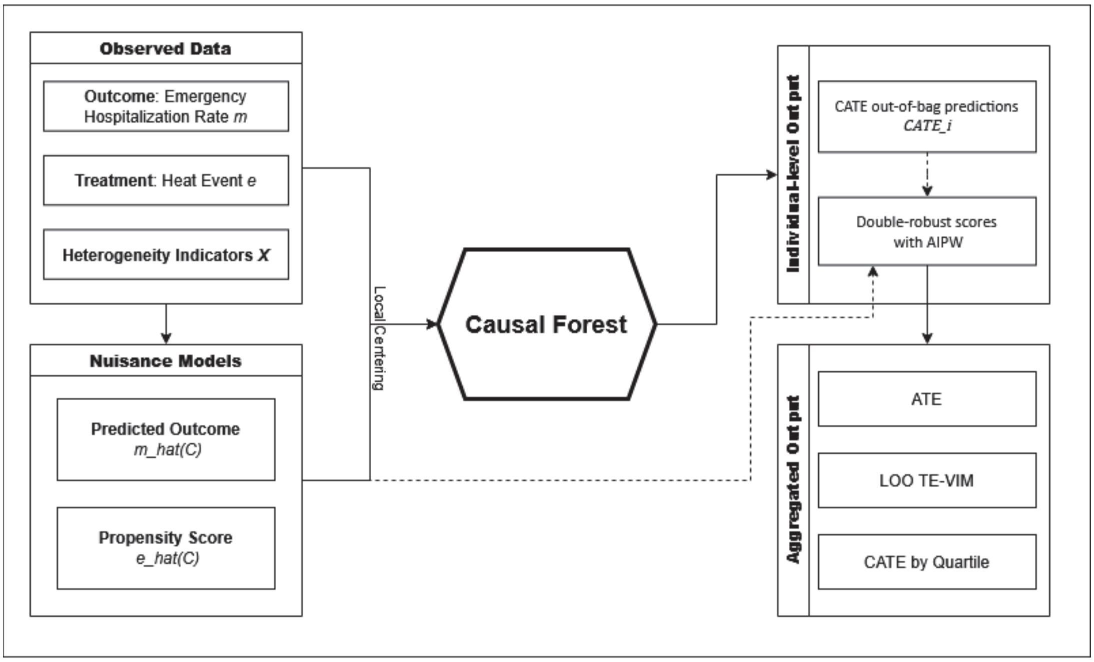


**Fig. S2**: Methodological workflow for generalized random forest. $\hat{m}$: Predicted emergency hospitalization rate given confounders (district, month, year). $\hat{e}$: Predicted probability of heat event given confounders. AIPW: augmented inverse probability weighting. Dashed arrow indicate that nuisance models and individual CATE predictions were used for constructing the doubly robust AIPW scores. LOO TE-VIM: Leave-one-out treatment effect variable importance metric.

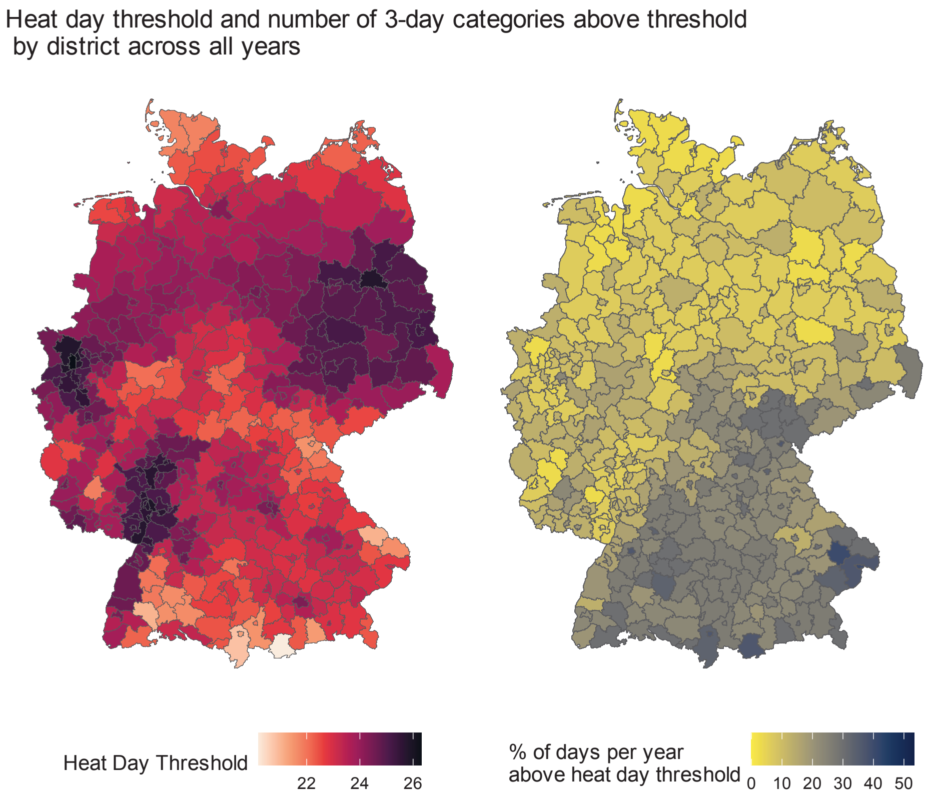


**Fig. S3**: 98th percentile max temperature threshold and number of days above threshold by district.

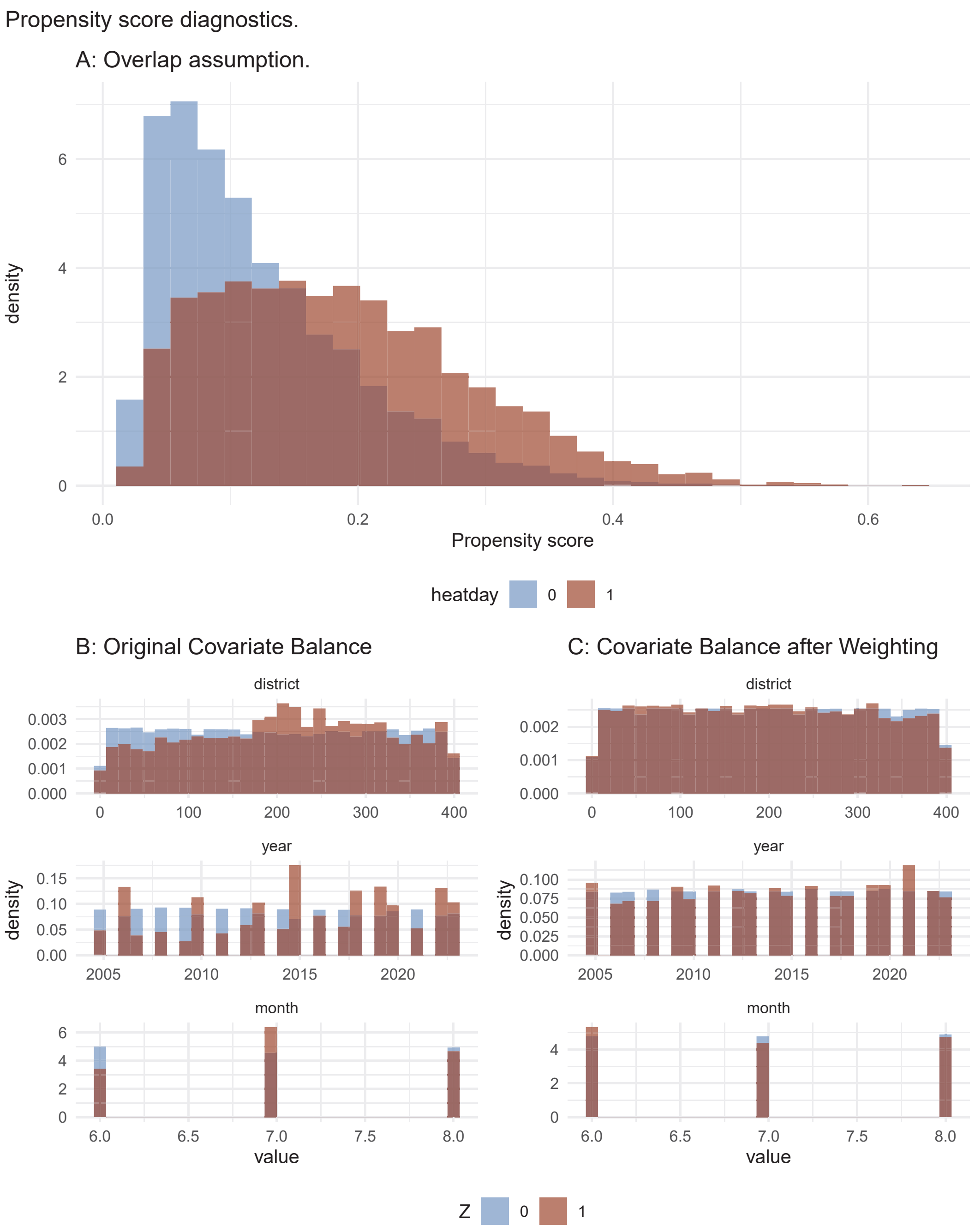


**Fig. S4**: Propensity score diagnostics. Overlap assumption and covariate balance before and after weighting with stabilized IPW.

#### Additional Analyses

##### Delayed Effects

**Table S3**: Calibration output of the grf.

| | Estimate | Std error | p-value |
|---|---|---|---|
| mean.forest.prediction | 1.143 | 0.289 | 3.832e-05 |
| differential.forest.prediction | 1.422 | 0.078 | <2.2e-16 |

A signficiant coefficient for the diferential forest predictions (p<2.2e-16) indicates that the causal forest sucessfully detected effect heterogeneity.

**Table S4**: Average Treatment Effects for main analysis, lags, heat definitions, and robustness check for excluding 2020-2023 from the data.

| | check | estimate | lower | upper |
|---|---|---|---|---|
| perc98th | -0.30 | -0.36 | -0.24 | |
| lag1 | -0.03 | -0.05 | -0.001 | |
| lag2 | -0.16 | -0.18 | -0.13 | |
| abs24 | -0.25 | -0.31 | -0.19 | |
| heatwave | -0.16 | -0.19 | -0.13 | |
| exclCovid | -0.32 | -0.38 | -0.27 | |

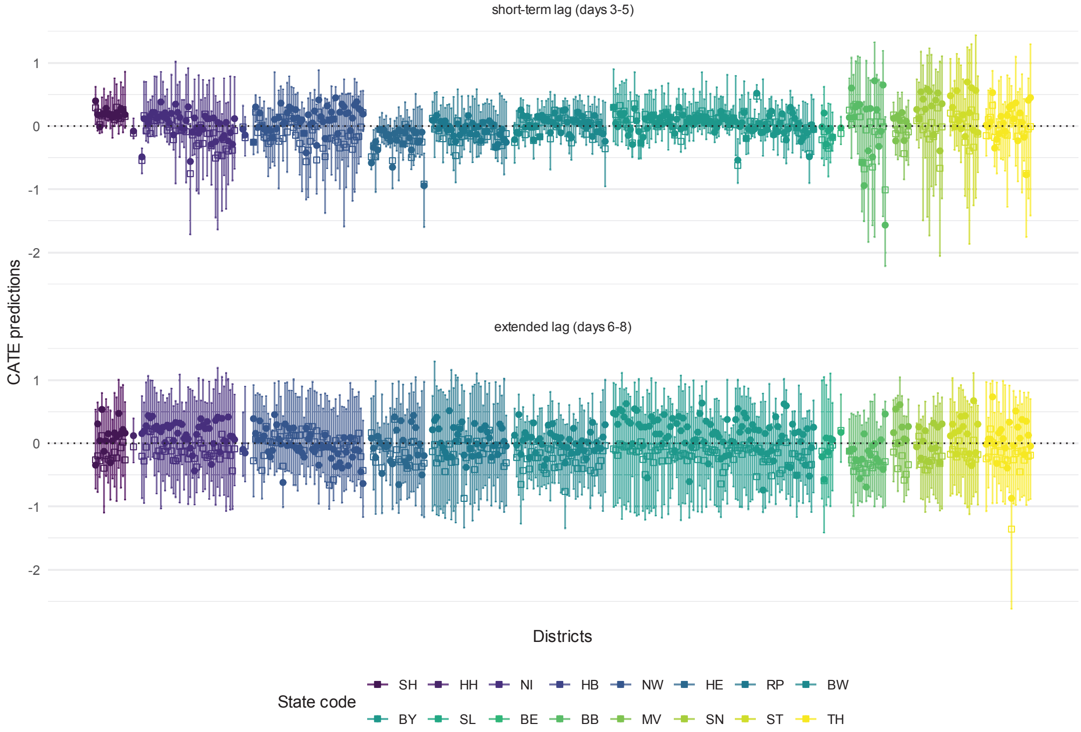


**Fig. S5**: Individual CATE predictions for delayed effect of a heat event (days 3-5 and 6-8) on the emergency hospitalization rate per 1,000, by district and grouped by state. Squares represent the mean, points the median. Bars represent the 25th to 75th percentile ranges.

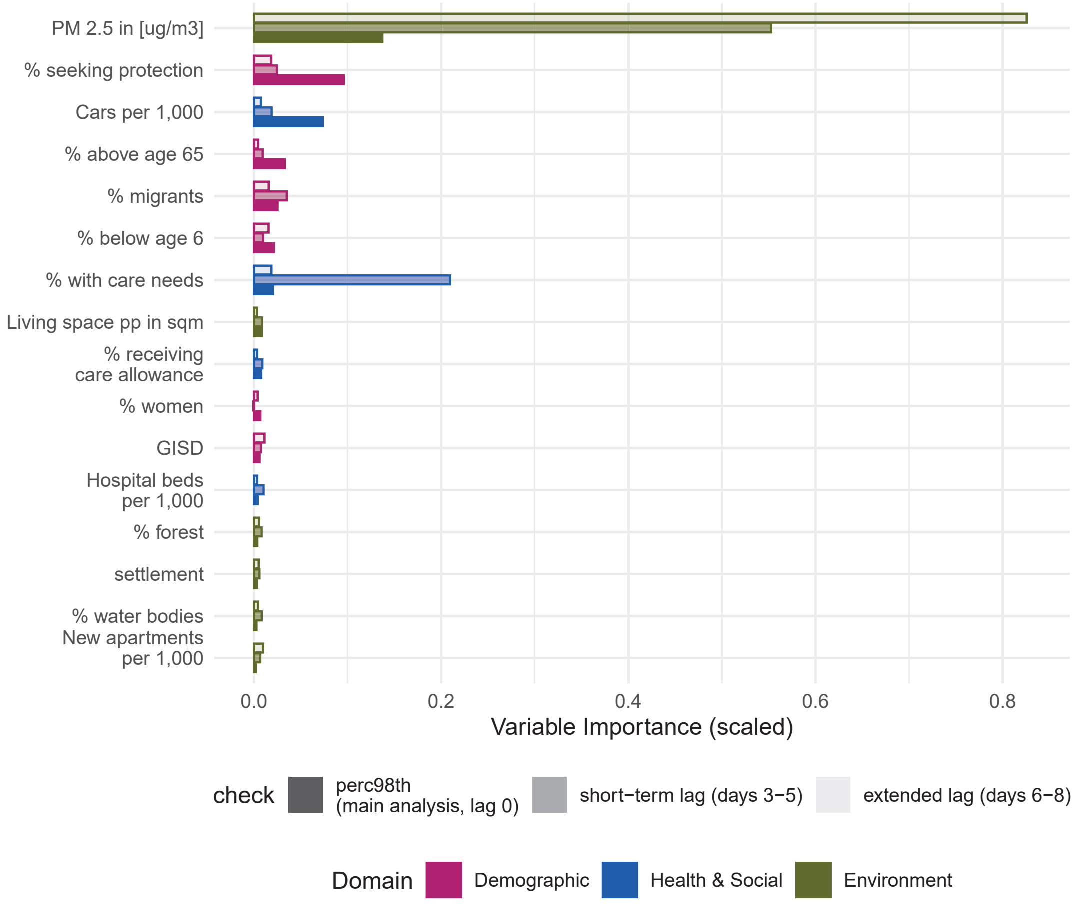


**Fig. S6**: Variable importance (scaled) for predicting effect heterogeneity in the immediate (days 0-2) and delayed effect (days 3-5 and 6-8) of a heat event on the emergency hospitalization rate by domain. Opaque bars represent the importance for the immediate response (main analysis).

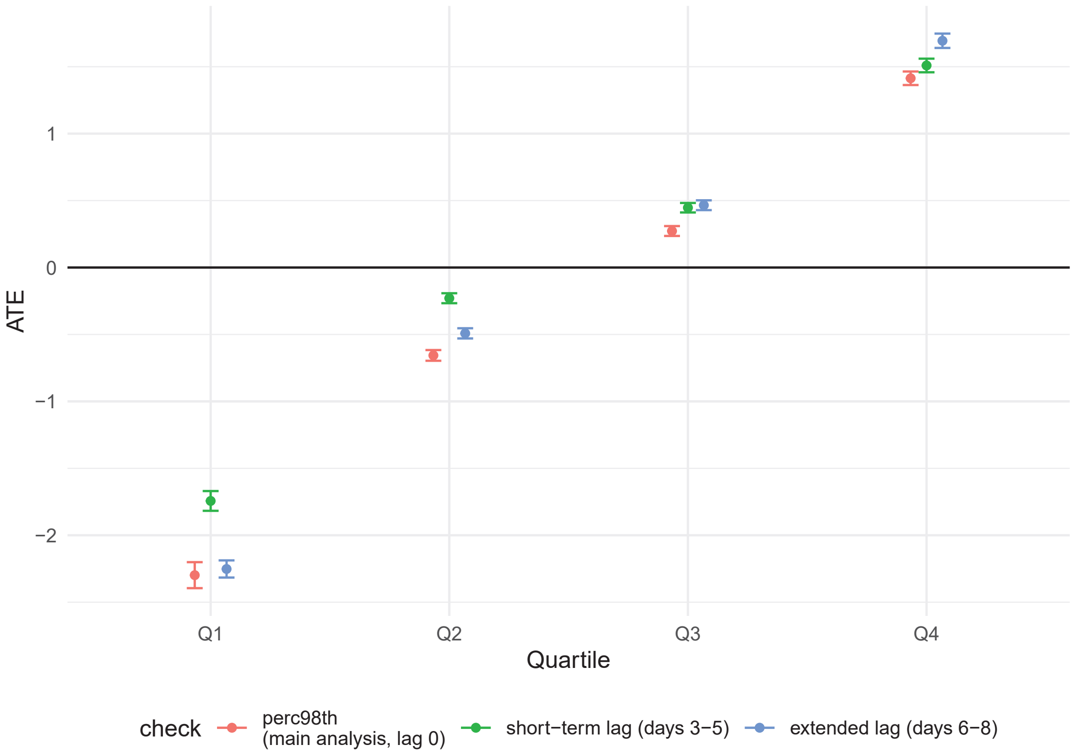


**Fig. S7**: Average treatment effect of a heat event on emergency hospitalization rate (95% CI) on days 3-5 and 6-8 since a heat event.

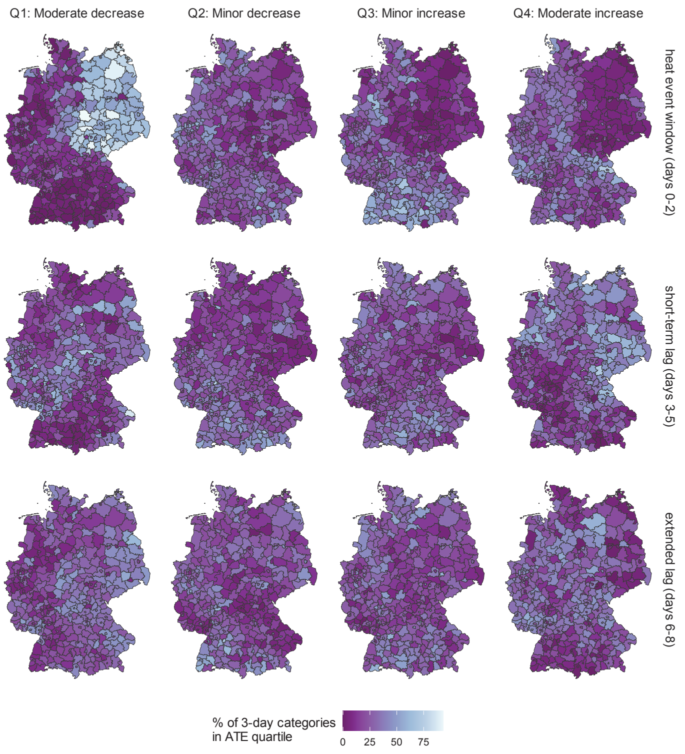


**Fig. S8**: Percentage of 3-day categories falling in a respective heat effect quartile by district across lags. The heat effect describes the immediate (lag 0, days 0-2) or delayed effect (days 3-5 or 6-8) of a heat event on the emergency hospitalization rate. Number in brackets shows the standard error.

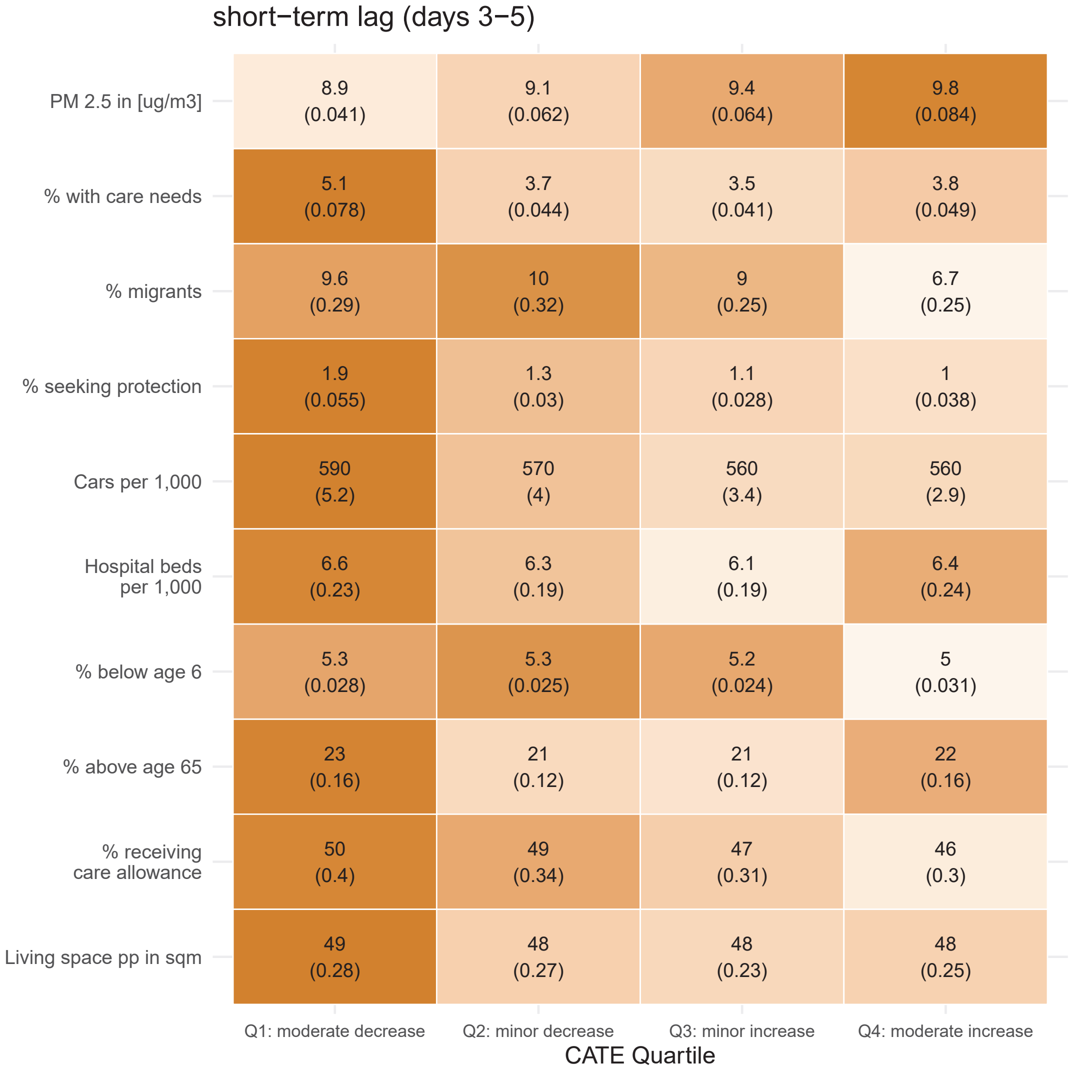


**Fig. S9**: Average indicator values within heat effect quartiles. Results are shown for the top 10 most important variables according to variable importance metric. The heat effect describes the delayed (days 3-5) effect of a heat event on the emergency hospitalization rate. Number in brackets shows the standard error. Darker colors indicate a larger average value of an indicator in a given quartile, in comparison to other quartiles.

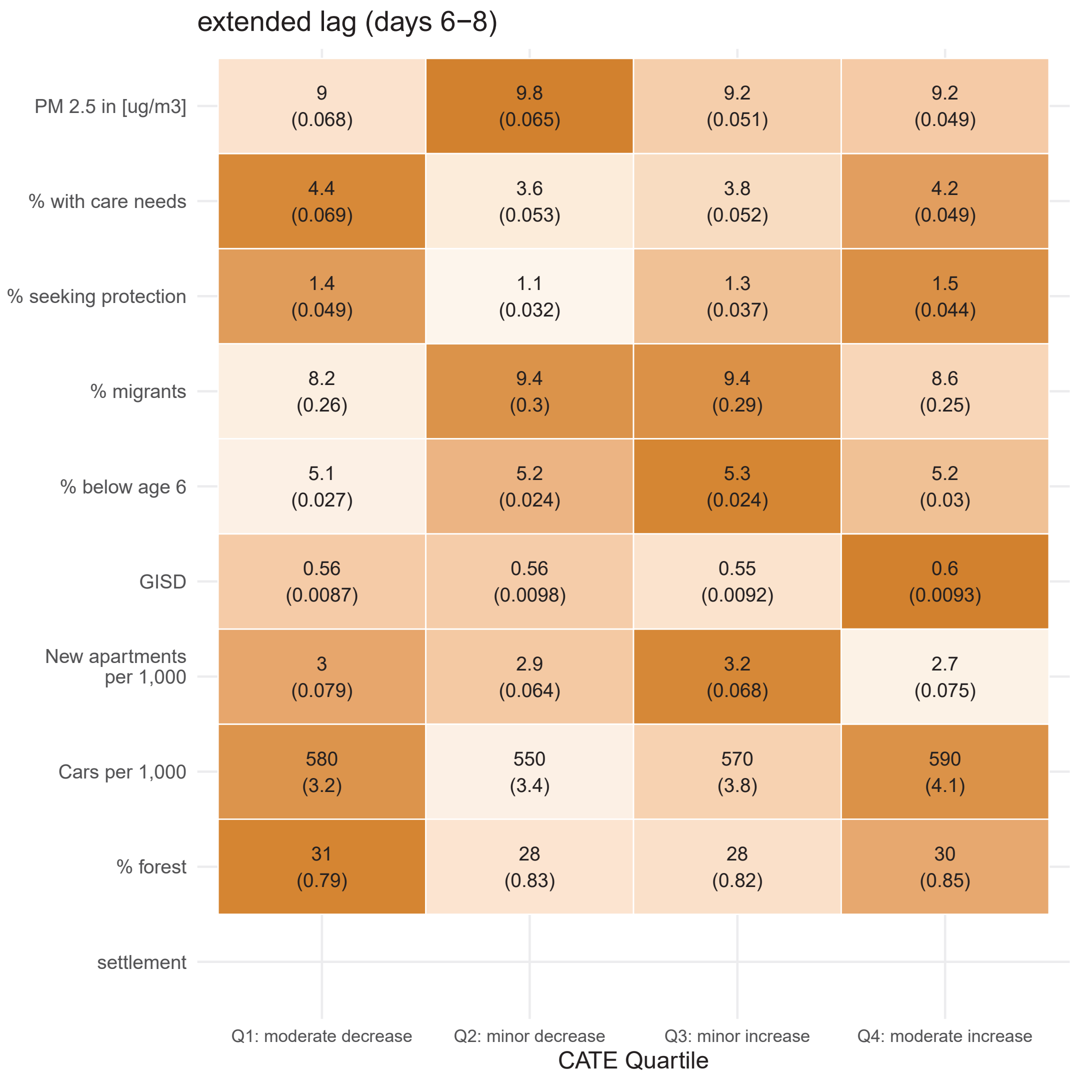


**Fig. S10**: Average indicator values within heat effect quartiles. Results are shown for the top 10 most important variables according to variable importance metric. The heat effect describes the delayed (days 6-8) effect of a heat event on the emergency hospitalization rate. Number in brackets shows the standard error. Darker colors indicate a larger average value of an indicator in a given quartile, in comparison to other quartiles.

#### Robustness Checks

We repeated our analysis with two other heat definitions (absolute cut-off and heatwave), and excluding the years of the Covid-19 pandemic. The ATE is comparable to the main results for the absolute cut-off and exclusion of 2020-2023 but we find a larger reduction in the emergency hospitalization rate of 0.13 (95%CI: -0.17;-0.10) per 1,000 for the heatwave definition (Table S4). CATE estimates across quartiles are similar across exposure definition, and Covid-19 sensitivity analysis for all quartiles except for Q1. The Q1 estimates are lower for the absolute heat cut-off and larger for the heatwave definition (Figure S11, (Figure S16). The quartile maps (Figure S13, (Figure S18) that show the most common CATE quartile are comparable for the absolute cut-off and Covid-19 robustness check. For the heatwave definition, we did not find a clear trend across regions with some districts in the East most often experiencing a moderate increase and some districts in the South and West experiencing a moderate increase in the emergency hospitalization rate. However, results based on the heatwave definition need to be interpreted with caution because the propensity score resulted in predicted heatwave probabilities of zero due to the low heatwave prevalence (6.22%) which might lead to unstable ATE estimates.

In regards to the drivers of effect heterogeneity, treatment effect heterogeneity for the absolute cut-off was explained mostly by the percentage of population with care needs but rankings were otherwise similar to the main analysis. For the heatwave definition, the effect modifiers of interest, especially air pollution and percentage of the population with care needs, explain a larger proportion of the treatment heterogeneity than for the main analysis (Figure S12. The covariate distribution differed for the heatwave definition, compared to the main analysis, with a larger percentage of people with care needs in the moderate decrease group rather than the moderate increase group(Figure S13,Figure S14).

When years 2020 to 2023 were excluded from the analysis, the percentage of people with care needs explained the largest percentage of effect heterogeneity instead of the percentage of people seeking protection compared to the main analysis. Otherwise, results are similar (Figure S17, Figure S19).

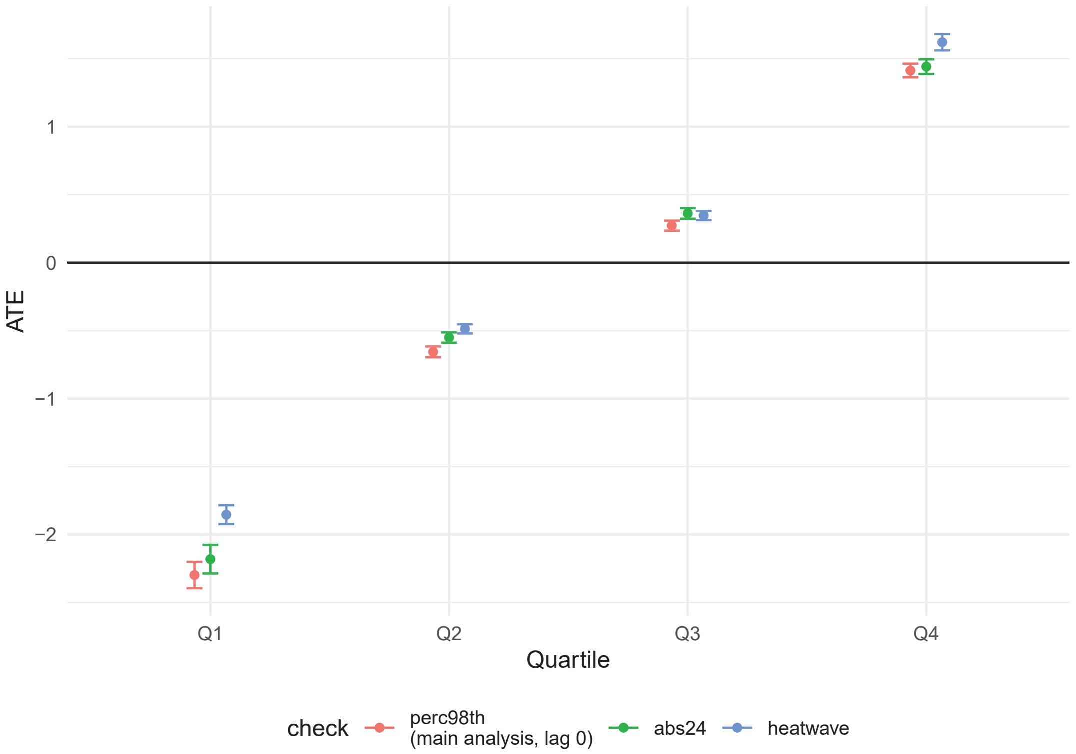


**Fig. S11**: Average treatment effect of a heat event on emergency hospitalization rate (95% CI) for different exposure definitions. Results based on the heatwave definition need to be interpreted with caution because the propensity score resulted in predicted probabilities for a heatwave of zero due to the low prevalence (6.22%) which might lead to unstable ATE estimates.

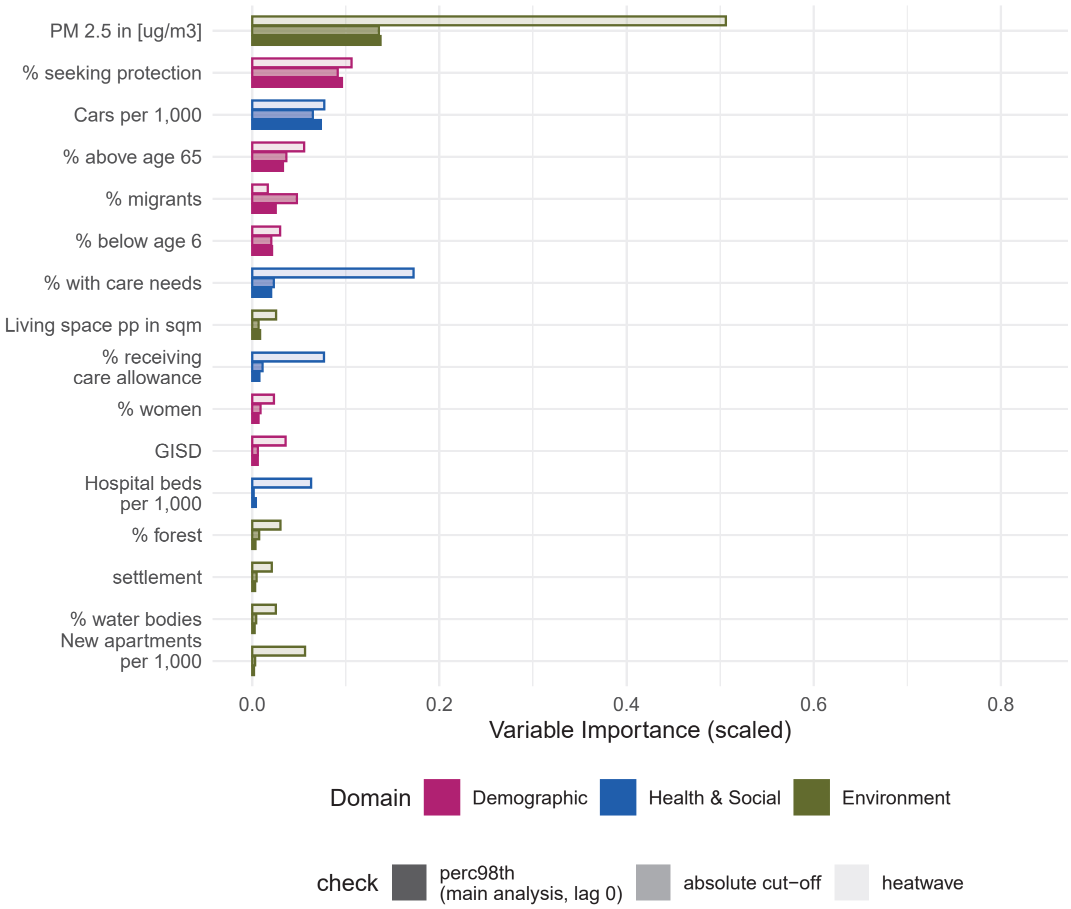


**Fig. S12**: Variable importance (scaled) for predicting effect heterogeneity in the immediate effect of a heat event on the emergency hospitalization rate by domain, across exposure definitions. Opaque bars represent the importance for the main analysis. Results based on the heatwave definition need to be interpreted with caution because the propensity score resulted in predicted probabilities for a heatwave of zero due to the low prevalence (6.22%) which might lead to unstable ATE estimates.

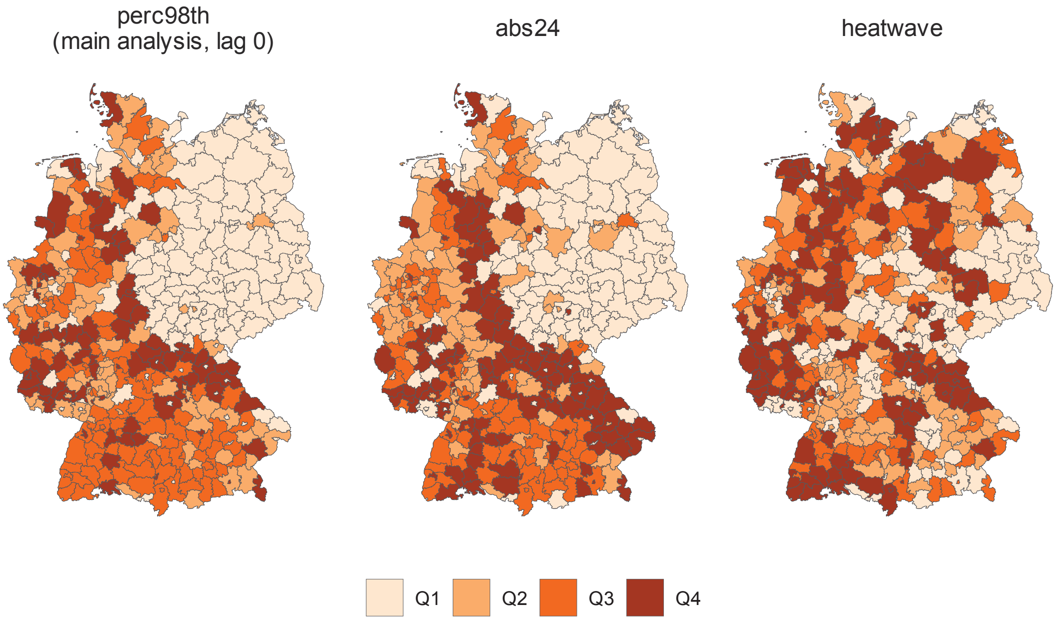


**Fig. S13**: Most common quartile of the immediate effect of a heat event on emergency hospitalizations across heat definitions, by district. Results based on the heatwave definition need to be interpreted with caution because the propensity score resulted in predicted probabilities for a heatwave of zero due to the low prevalence (6.22%) which might lead to unstable ATE estimates.

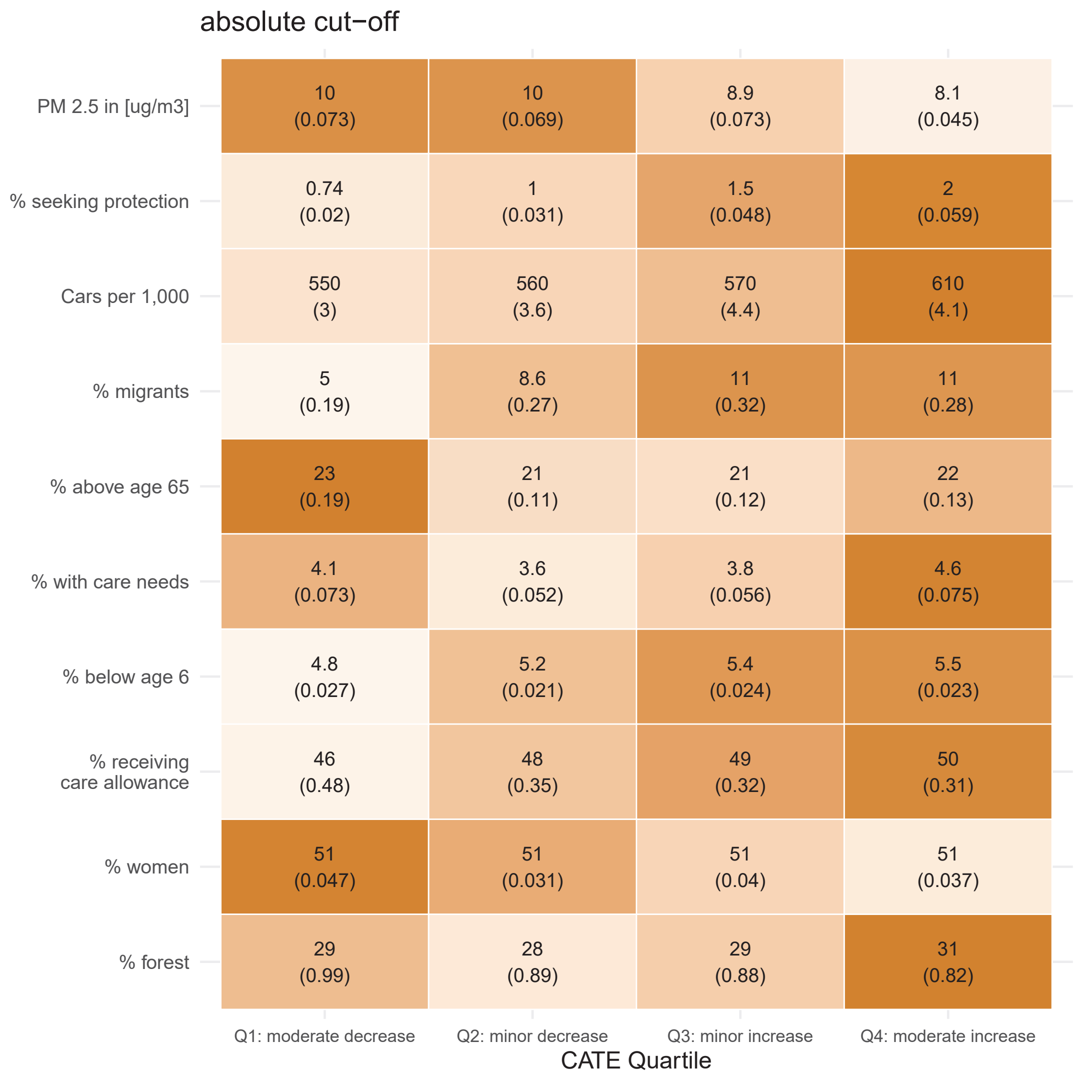


**Fig. S14**: Average indicator values within heat effect quartiles for absolute heat cut-off. Results are shown for the top 10 most important variables according to variable importance metric. The heat effect describes the immediate effect of a heat event on the emergency hospitalization rate. Number in brackets shows the standard error. Darker colors indicate a larger average value of an indicator in a given quartile, in comparison to other quartiles.

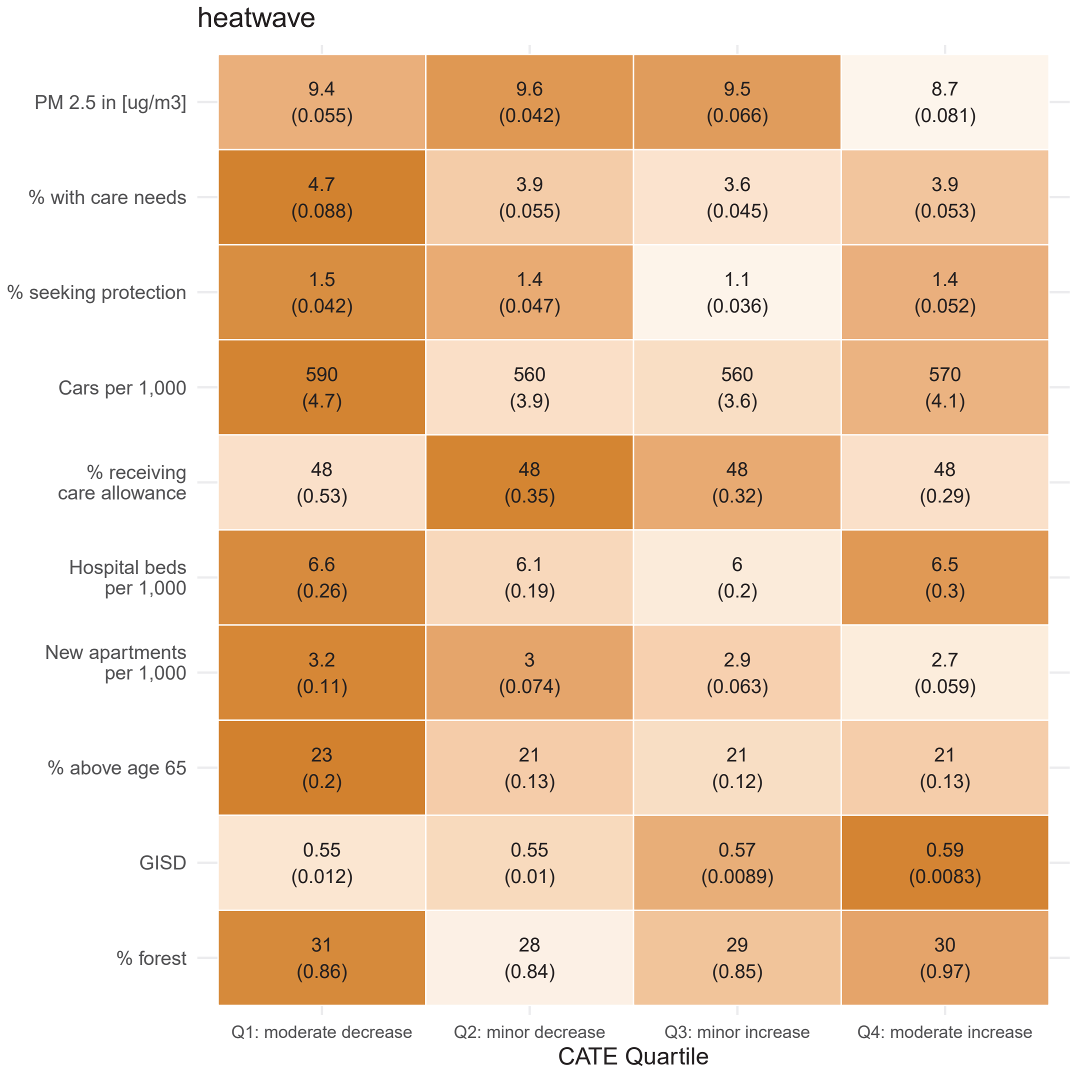


**Fig. S15**: Average indicator values within heat effect quartiles for heatwave definition. Results are shown for the top 10 most important variables according to variable importance metric. The heat effect describes the immediate effect of a heat event on the emergency hospitalization rate. Number in brackets shows the standard error. Darker colors indicate a larger average value of an indicator in a given quartile, in comparison to other quartiles. Results based on the heatwave definition need to be interpreted with caution because the propensity score resulted in predicted probabilities for a heatwave of zero due to the low prevalence (6.22%) which might lead to unstable ATE estimates.

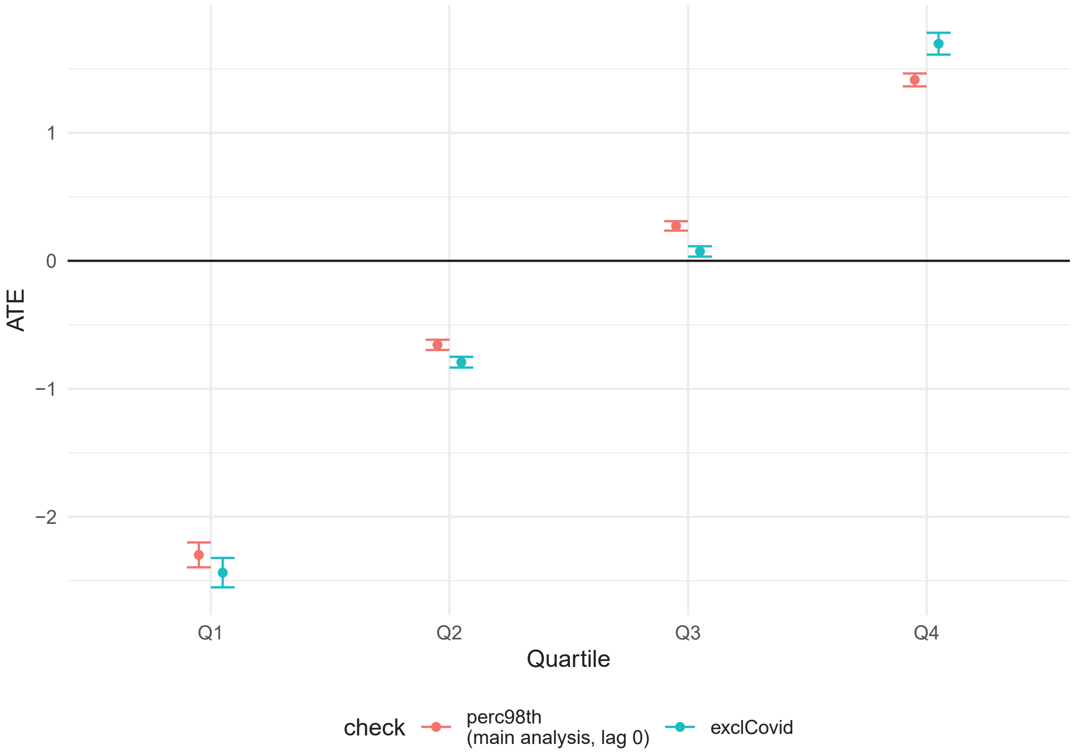


**Fig. S16**: Average treatment effect of a heat event on emergency hospitalization rate (95% CI) for main analysis compared to excluding Covid-19 years.

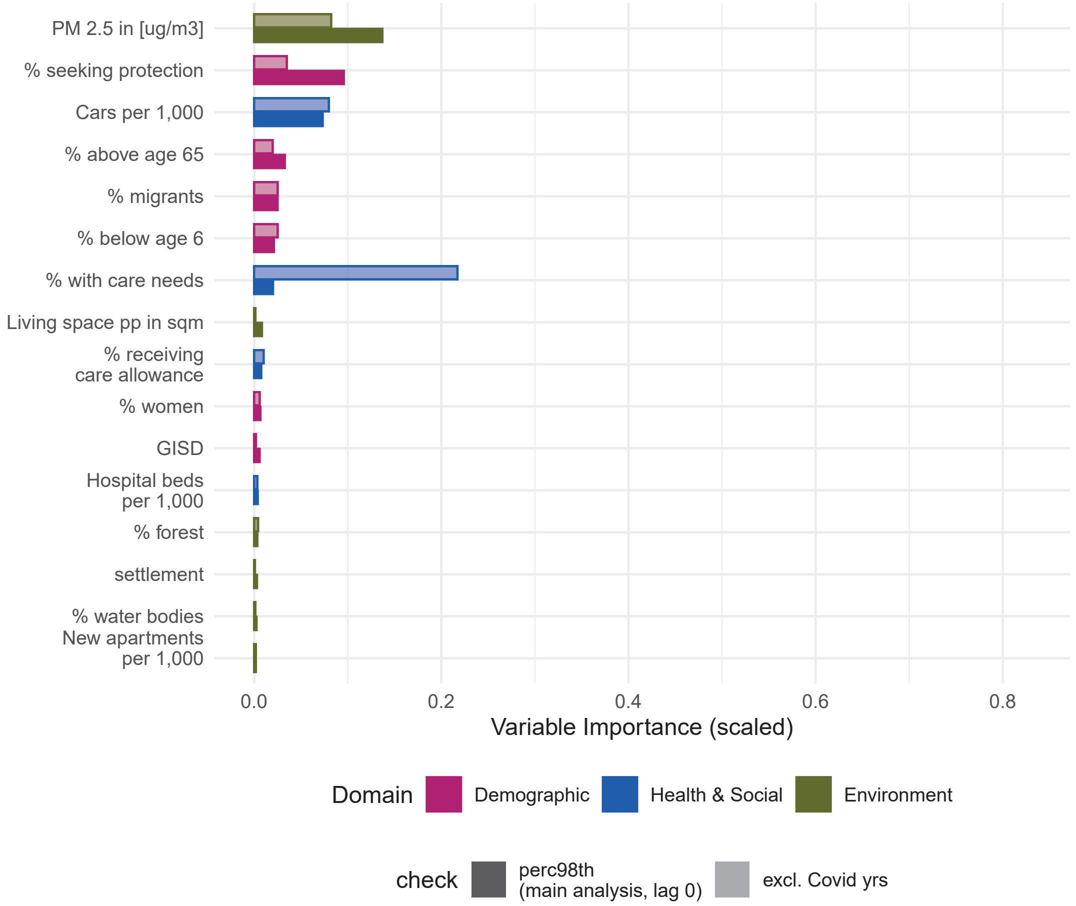


**Fig. S17**: Variable importance (scaled) for predicting effect heterogeneity in the immediate effect of a heat event on the emergency hospitalization rate by domain. Comparing main analysis with excluding Covid-19 years (2020-2023). Opaque bars represent the importance for the main analysis.

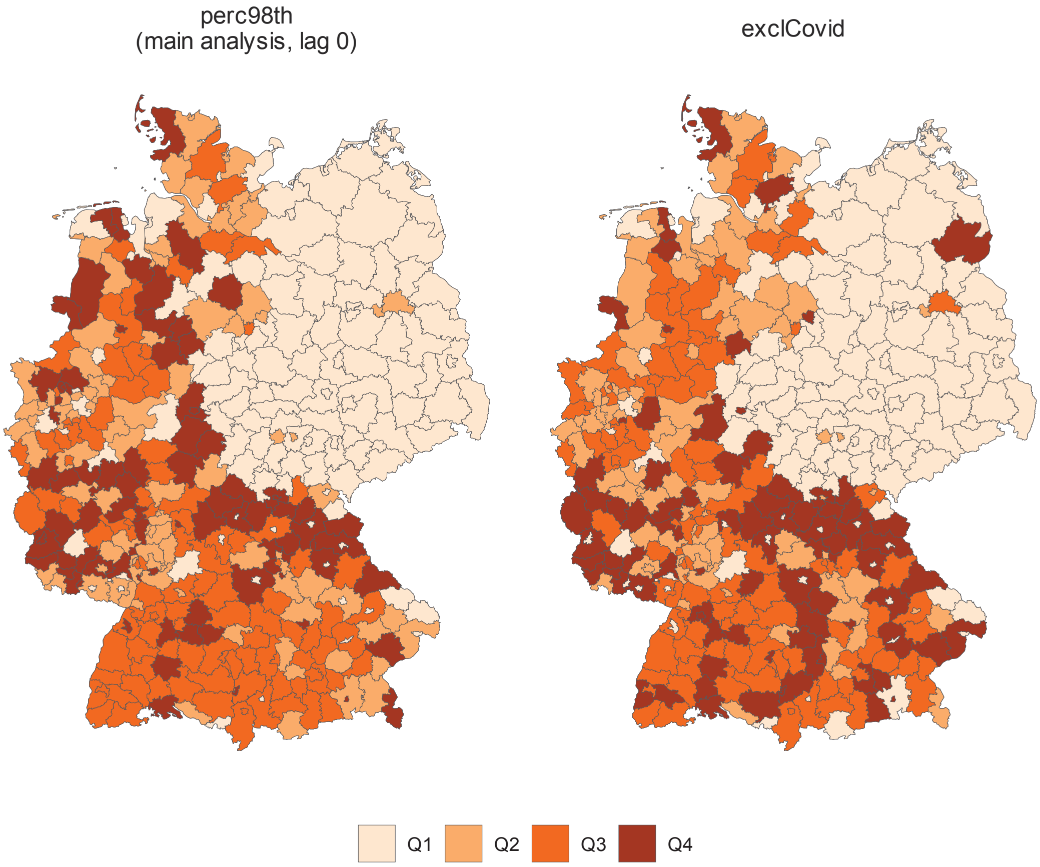


**Fig. S18**: Most common quartile of the immediate effect of a heat event on emergency hospitalizations by district, comparing main analysis and excluding Covid-19 years (2020-2023).

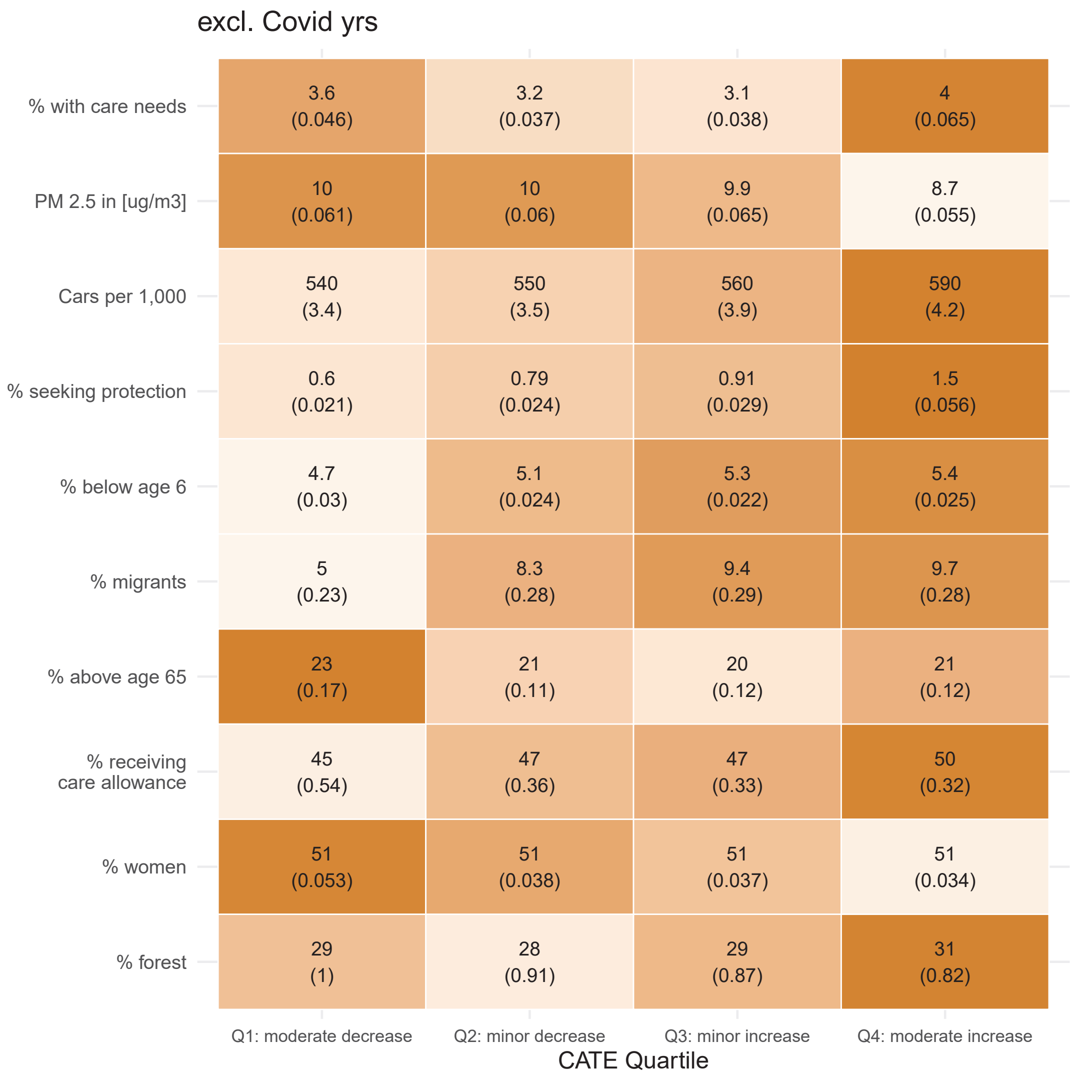


**Fig. S19**: Average indicator values within heat effect quartiles for excluding Covid-19 years (2020-2023). Results are shown for the top 10 most important variables according to variable importance metric. The heat effect describes the immediate effect of a heat event on the emergency hospitalization rate. Number in brackets shows the standard error. Darker colors indicate a larger average value of an indicator in a given quartile, in comparison to other quartiles.

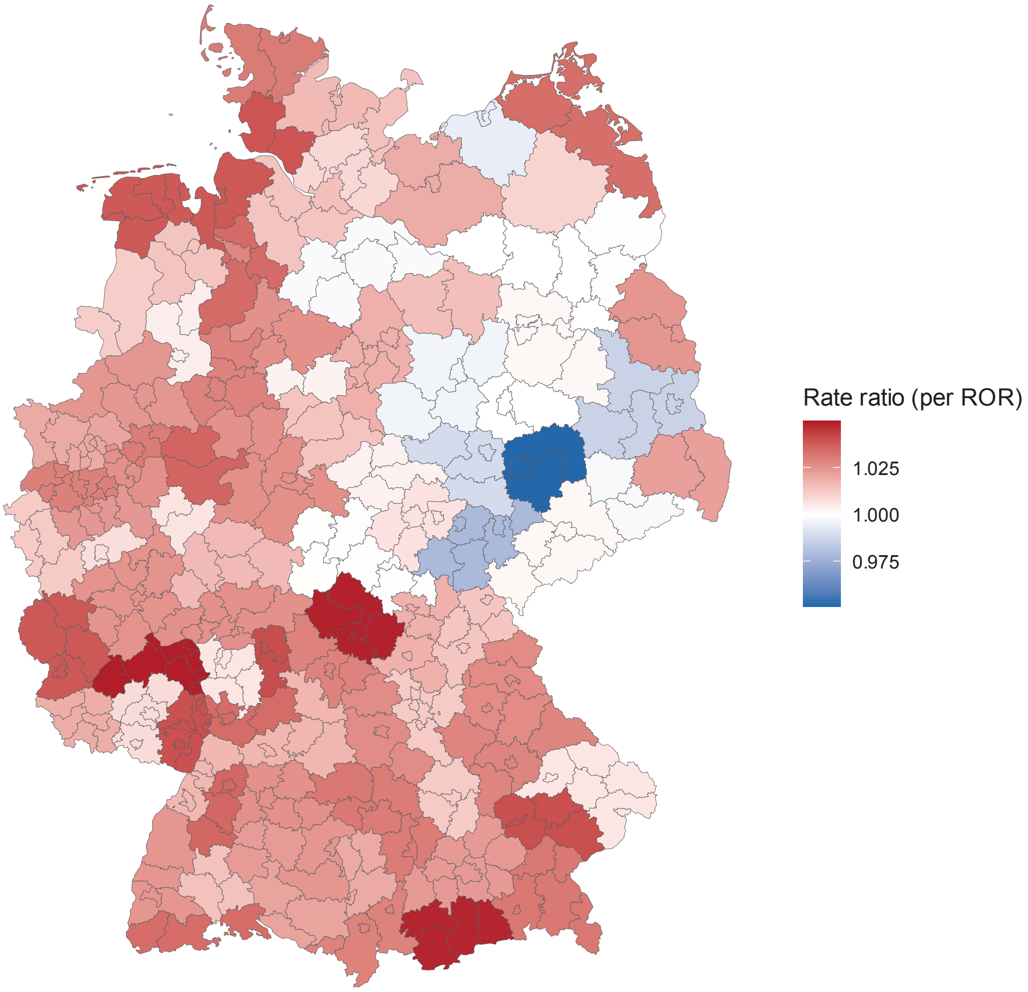


**Fig. S20**: Exponentiated coefficient for heat event-emergency hospitalization association. Coefficients were obtained from a simple Poisson regression model, controlled for year and month and including a population offset. Model was fit separately for each of the 96 ”Raumordnungsregionen” (ROR, granularity lies between NUTS-2 and NUTS-3) instead of 400 districts to reduce noise.

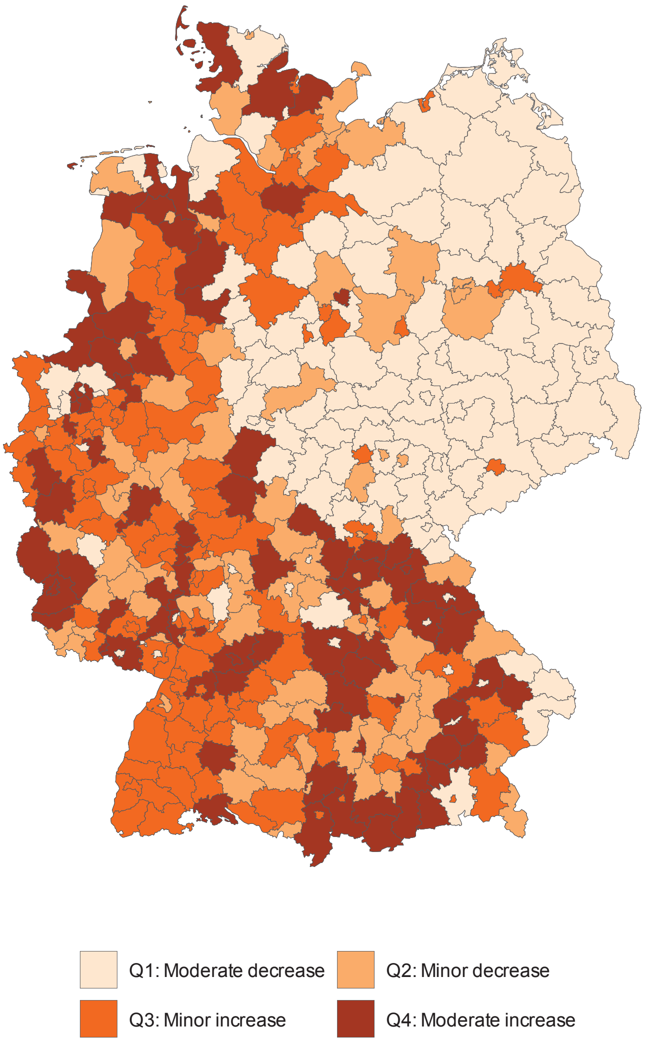


**Fig. S21**: Most common quartile of the effect of a heat event on emergency hospitalizations by district based on a grf that was fit without indicators that show a gradient along the former East-West border (% population above 65 and below 6, German Index of Socioeconomic Deprivation, % migrants, living space) .

TC:endignore

### References

[1] Guo C, Ge E, Lee S, Lu Y, Bassill NP, Zhang N, et al. Impact of heat on emergency hospital admission in Texas: geographic and racial/ethnic disparities. Journal of exposure science & environmental epidemiology. 2024 11;34(6):927–934. https://doi.org/10.1038/S41370-023-00590-6.

[2] Chen C, Schwarz L, Rosenthal N, Marlier ME, Benmarhnia T. Exploring spatial heterogeneity in synergistic effects of compound climate hazards: Extreme heat and wildfire smoke on cardiorespiratory hospitalizations in California. Science advances. 2024 2;10(5). https://doi.org/10.1126/SCIADV.ADJ7264.

[3] Patel D, Jian L, Xiao J, Jansz J, Yun G, Robertson A. Joint effect of heatwaves and air quality on emergency department attendances for vulnerable population in

Perth, Western Australia, 2006 to 2015. Environmental research. 2019 7;174:80–87. https://doi.org/10.1016/J.ENVRES.2019.04.013.

[4] Anderko L, Davies-Cole J, Strunk A. Identifying populations at risk: interdisciplinary environmental climate change tracking. Public health nursing (Boston, Mass). 2014 11;31(6):484–491. https://doi.org/10.1111/PHN.12164.

[5] Clark A, Grineski S, Curtis DS, Cheung ESL. Identifying groups at-risk to extreme heat: Intersections of age, race/ethnicity, and socioeconomic status. Environment international. 2024 9;191. https://doi.org/10.1016/J.ENVINT.2024.108988.

[6] Thomson TN, Rupasinghe R, Hennessy D, Easton M, Stewart T, Mulvenna V. Population vulnerability to heat: A case-crossover analysis of heat health alerts and hospital morbidity data in Victoria, Australia. Australian and New Zealand journal of public health. 2023 12;47(6). https://doi.org/10.1016/J.ANZJPH.2023.100092.

[7] Salvador C, Gullón P, Franco M, Vicedo-Cabrera AM. Heat-related first cardiovascular event incidence in the city of Madrid (Spain): Vulnerability assessment by demographic, socioeconomic, and health indicators. Environmental research. 2023 6;226. https://doi.org/10.1016/J.ENVRES.2023.115698.

[8] Guo C, Ge E, Yu M, Li C, Lao X, Li S, et al. Impact of heat on emergency hospital admissions related to kidney diseases in Texas: Uncovering racial disparities. The Science of the total environment. 2024 1;909. https://doi.org/10.1016/J.SCITOTENV.2023.168377.

[9] Gronlund CJ. Racial and socioeconomic disparities in heat-related health effects and their mechanisms: a review. Current epidemiology reports. 2014 9;1(3):165–173. https://doi.org/10.1007/S40471-014-0014-4.

[10] Graffy PM, Barrett BW, Horton DE, Kho AN, Allen NB. Critical Temperature Thresholds for Identifying Vulnerability to Heat-Related Excess Cardiovascular Morbidity and Mortality. Journal of the American Heart Association. 2026 1;15(2). https://doi.org/10.1161/JAHA.125.046117.

[11] Hajat S, Kosatky T. Heat-related mortality: a review and exploration of heterogeneity. Journal of Epidemiology & Community Health. 2010 9;64(9):753–760. https://doi.org/10.1136/JECH.2009.087999.

[12] Christenson M, Geiger SD, Phillips J, Anderson B, Losurdo G, Anderson HA. Heat Vulnerability Index Mapping for Milwaukee and Wisconsin. Journal of public health management and practice : JPHMP. 2017;23(4):396–403. https://doi.org/10.1097/PHH.0000000000000352.

[13] Rizmie D, de Preux L, Miraldo M, Atun R. Impact of extreme temperatures on emergency hospital admissions by age and socio-economic deprivation in England. Social Science & Medicine. 2022 9;308:115193. https://doi.org/10.1016/J.SOCSCIMED.2022.115193.

[14] Xiao J, Spicer T, Jian L, Yun GY, Shao C, Nairn J, et al. Variation in Population Vulnerability to Heat Wave in Western Australia. Frontiers in public health. 2017 4;5(APR). https://doi.org/10.3389/FPUBH.2017.00064.

[15] Arsad FS, Hod R, Ahmad N, Ismail R, Mohamed N, Baharom M, et al. The Impact of Heatwaves on Mortality and Morbidity and the Associated Vulnerability Factors: A Systematic Review. International journal of environmental research

and public health. 2022 12;19(23). https://doi.org/10.3390/IJERPH192316356.

[16] Wu WJ, Hutton J, Zordan R, Ranse J, Crilly J, Tutticci N, et al. Review article: Scoping review of the characteristics and outcomes of adults presenting to the emergency department during heatwaves. Emergency medicine Australasia : EMA. 2023 12;35(6):903–920. https://doi.org/10.1111/1742-6723.14317.

[17] Adnan Bukhari H. A Systematic Review on Outcomes of Patients with Heatstroke and Heat Exhaustion. Open access emergency medicine : OAEM. 2023;15:343–354. https://doi.org/10.2147/OAEM.S419028.

[18] Li M, Gu S, Bi P, Yang J, Liu Q. Heat Waves and Morbidity: Current Knowledge and Further Direction-A Comprehensive Literature Review. International Journal of Environmental Research and Public Health. 2015 may;12(5):5256–5283. https://doi.org/10.3390/IJERPH120505256.

[19] Hansen A, Bi P, Nitschke M, Pisaniello D, Newbury J, Kitson A. Older persons and heat-susceptibility: the role of health promotion in a changing climate. Health promotion journal of Australia : official journal of Australian Association of Health Promotion Professionals. 2011;22 Spec No. https://doi.org/10.1071/HE11417.

[20] Brammer M, Gerstner D, Heinze S, Grümme L, Kneißl K, Trentzsch H, et al. City characteristics and heat vulnerability: insights from emergency medical services in Bavaria, Germany. International journal of biometeorology. 2026 2;70(2):35. https://doi.org/10.1007/S00484-025-03076-2.

[21] Yang C, Li X, Zhang W, Tao Y, Zhu P, Lu C, et al. Associations between apparent temperatures and emergency ambulance calls in Wuxi, China: a time series analysis. Frontiers in public health. 2025;13. https://doi.org/10.3389/FPUBH.2025.1652961.

[22] Hajdu T. Heterogeneous impacts of climate change on morbidity. Economics and human biology. 2025 9;58. https://doi.org/10.1016/J.EHB.2025.101517.

[23] Niu L, Girma B, Liu B, Schinasi LH, Clougherty JE, Sheffield P. Temperature and mental health-related emergency department and hospital encounters among children, adolescents and young adults. Epidemiology and psychiatric sciences. 2023 4;32. https://doi.org/10.1017/S2045796023000161.

[24] Kovats RS, Hajat S. Heat stress and public health: A critical review. Annual Review of Public Health. 2008 apr;29(Volume 29, 2008):41–55. https://doi.org/10.1146/annurev.publhealth.29.020907.090843.

[25] Liao S, Pan W, Wen L, Chen R, Pan D, Wang R, et al. Temperature-related hospitalization burden under climate change. Nature. 2025 8;644(8078):960–968. https://doi.org/10.1038/S41586-025-09352-W.

[26] Ulrich SE, Sugg MM, Roy M, Runkle JD. Temperature extremes and maternal health: differential risks of severe maternal morbidity during heatwaves and coldwaves in North Carolina. International journal of biometeorology. 2026 2;70(2). https://doi.org/10.1007/S00484-025-03079-Z.

[27] Schwarz L, Castillo EM, Chan TC, Brennan JJ, Sbiroli ES, Carrasco-Escobar G, et al. Heat Waves and Emergency Department Visits Among the Homeless, San Diego, 2012-2019. American journal of public health. 2022 1;112(1):98–106. https://doi.org/10.2105/AJPH.2021.306557.

[28] Lavigne E, Maltby A, Côté JN, Weinberger KR, Hebbern C, Vicedo-Cabrera AM, et al. The effect modification of extreme temperatures on mental and behavior disorders by environmental factors and individual-level characteristics in Canada. Environmental research. 2023 2;219. https://doi.org/10.1016/J.ENVRES.2022.114999.

[29] Bai L, Woodward A, Cirendunzhu, Liu Q. County-level heat vulnerability of urban and rural residents in Tibet, China. Environmental Health. 2016 1;15(1):3. https://doi.org/10.1186/S12940-015-0081-0.

[30] Yu J, Castellani K, Forysinski K, Gustafson P, Lu J, Peterson E, et al. Geospatial indicators of exposure, sensitivity, and adaptive capacity to assess neighbourhood variation in vulnerability to climate change-related health hazards. Environmental Health. 2021 12;20(1):31. https://doi.org/10.1186/S12940-021-00708-Z.

[31] Xu R, Zhao Q, Coelho MSZS, Saldiva PHN, Abramson MJ, Li S, et al. Socioeconomic inequality in vulnerability to all-cause and cause-specific hospitalisation associated with temperature variability: a time-series study in 1814 Brazilian cities. The Lancet Planetary health. 2020 12;4(12):e566–e576. https://doi.org/10.1016/S2542-5196(20)30251-5.

[32] Lung SCC, Yeh JCJ, Hwang JS, Chen LS. Evaluation of heat warning thresholds with multiple lagged and cumulative health impacts based on a 20-year population database. Scientific reports. 2026 12;16(1). https://doi.org/10.1038/S41598-025-31832-2.

[33] Meadows J, Mansour A, Gatto MR, Li A, Howard A, Bentley R. Mental illness and increased vulnerability to negative health effects from extreme heat events: a systematic review. Psychiatry research. 2024 2;332. https://doi.org/10.1016/J.PSYCHRES.2023.115678.

[34] Hammer J, Ruggieri DG, Thomas C, Caum J. Local Extreme Heat Planning: an Interactive Tool to Examine a Heat Vulnerability Index for Philadelphia, Pennsylvania. Journal of Urban Health : Bulletin of the New York Academy of Medicine. 2020 8;97(4):519. https://doi.org/10.1007/S11524-020-00443-9.

[35] Byun G, Kim S, Festa N, Choi Y, Lee WW, Lee JT, et al. Effects of ambient temperature on hospital admissions and mortality among older adults with and without dementia in South Korea. International journal of epidemiology. 2025 8;54(4). https://doi.org/10.1093/IJE/DYAF142.

[36] Kim S, Byun G, Lee JT. Association between non-optimal temperature and cardiovascular hospitalization and its temporal variation at the intersection of disability. The Science of the total environment. 2023 12;904. https://doi.org/10.1016/J.SCITOTENV.2023.166874.

[37] Requia WJ, Hoinaski L, Yang J, Adams MD, Yazdi MD, Rodrigues da Silva Júnior FM, et al. The role of green areas in modifying heat-related circulatory and respiratory hospital admissions in Brazil. Environment international. 2025 9;203. https://doi.org/10.1016/J.ENVINT.2025.109791.

[38] Kim EJ, Kim H. Effect modification of individual- and regional-scale characteristics on heat wave-related mortality rates between 2009 and 2012 in Seoul, South Korea. The Science of the total environment. 2017 10;595:141–148. https://doi.org/10.1016/J.SCITOTENV.2017.03.248.

[39] Wong MS, Ho HC, Tse A. Geospatial context of social and environmental factors associated with health risk during temperature extremes: Review and discussion.

Geospatial health. 2020;15(1):168–173. https://doi.org/10.4081/GH.2020.814.

[40] Ma Y, Chen C, Aguilera R, Gershunov A, Jerrett M, Connolly R, et al. Spatial heterogeneity in synergistic effects of extreme heat and NO2 exposures on cardiorespiratory hospitalizations in California. Epidemiology (Cambridge, Mass). 2026 3;https://doi.org/10.1097/EDE.0000000000001970.

[41] Badaloni C, De Sario M, Caranci N, de' Donato F, Bolignano A, Davoli M, et al. A spatial indicator of environmental and climatic vulnerability in Rome. Environment International. 2023 6;176:107970. https://doi.org/10.1016/J.ENVINT.2023.107970.

[42] Rastogi D, Christian J, Tuccillo J, Christian B, Kapadia AJ, Hanson HA. Exploring the Spatial Patterning of Sociodemographic Disparities in Extreme Heat Exposure at Multiple Scales Across the Conterminous United States. GeoHealth. 2023 10;7(10). https://doi.org/10.1029/2023GH000864.

[43] Fastl C, Arnberger A, Gallistl V, Stein VK, Dorner TE. Heat vulnerability: health impacts of heat on older people in urban and rural areas in Europe. Wiener klinische Wochenschrift. 2024 9;136(17-18):507–514. https://doi.org/10.1007/S00508-024-02419-0.

[44] Atmospheric Composition Analysis Group (ACAG).: Satellite-derived PM2.5 (V6.GL.02.04). Atmospheric Composition Analysis Group (ACAG). Available from: https://sites.wustl.edu/acag/surface-pm2-5/#V6.GL.02.04.

[45] Cornes RC, van der Schrier G, van den Besselaar EJM, Jones PD. An Ensemble Version of the E-OBS Temperature and Precipitation Data Sets. Journal of Geophysical Research: Atmospheres. 2018 9;123(17):9391–9409. https://doi.org/https://doi.org/10.1029/2017JD028200.
